# Anticholinergic Medications and Health in Adults with Autism or Intellectual Disability

**DOI:** 10.64898/2026.10.26.26366518

**Authors:** Goldie A. McQuaid, Sha Tao, Lindsay L. Shea, Gregory L. Wallace

## Abstract

**Objective:** Contributors to autistic adults’ elevated risk for negative aging-related outcomes, including Alzheimer’s disease and related dementias (ADRD), falls, and mortality remain underexplored. One candidate factor is exposure to medications with anticholinergic (AC) effects. Thus, this study assessed prevalence of medications with AC effects, and their associations with ADRD, falls, and mortality across four groups: autism-only, autism+intellectual disability (ID), ID-only, and controls.

**Methods:** We conducted a retrospective cohort study of 2017-2019 US linked Medicare and Medicaid claims data for 40+ year-olds. Autism, ID, ADRD, and falls were identified using validated algorithms, and mortality was based on Medicare/Medicaid death indicators. Medications were coded using a comprehensive AC potency scale. AC medication exposure effects and interactions between AC potency and group were examined using modified Poisson regressions. Sensitivity analyses to address indication bias excluded persons prescribed antidepressant and/or antipsychotic medication(s).

**Results:** All three clinical groups experienced significantly greater AC medication exposure (autism+ID: 92.5%>ID-only: 88.6%>autism-only: 80.0%) than non-autistic/non-ID controls (61.3%). Additionally, AC medications were more frequently associated with ADRD, falls, and mortality in all of these groups. Stepwise associations between AC potency levels and ADRD, falls, and mortality were found, and these effects were more prominent for persons with autism and/or ID relative to the control sample.

**Interpretation:** This study provides a foundational examination of exposure to AC medication effects and their associations with aging-related outcomes in middle-age and older autistic adults and adults with ID. AC medication exposure represents one potential modifiable risk factor for negative aging-related outcomes in these groups.

## Introduction

Relatively little is known about aging among autistic people^1^; however, extant evidence indicates suboptimal outcomes. Notably, elevated rates of Alzheimer’s disease and related dementias (ADRD) have been reported for autistic adults aged ≥65 years enrolled in Medicare, with a four-fold increase in ADRD compared to general population Medicare enrollees.^2^ Two subsequent studies, one of Medicaid enrollees, and the other including linked Medicare and Medicaid data, found elevated prevalence and incidence of ADRD among autistic adults <65 years^3^ and ≥65 years.^4^ Examining aging-related outcomes beyond ADRD, a Swedish population-based longitudinal study found that autistic adults ≥45 years experienced higher falls risk relative to non-autistic adults.^5^ Among Medicare enrollees in the US, autistic adults aged 65+ had higher rates of falls and fall-related injuries and hospitalizations relative to non-autistic adults.^6^ Autistic adults also appear to face augmented mortality risk,^7,8^ including premature mortality,^9^ compared to non-autistic adults. Among Medicare-enrolled individuals 65+ years, autistic adults had a 2.87 times greater rate of mortality compared to non-autistic adults.^10^

Factors related to these increased risks for poorer cognitive and physical outcomes among autistic adults remain underexplored. One candidate factor meriting scrutiny is exposure to medications with anticholinergic (AC) effects. In the general aging population, exposure to medications with AC effects (including medications prescribed for their AC therapeutic properties and medications not prescribed for these properties but possessing AC effects^11^) are associated with negative outcomes, including cognitive impairments.^12^ More specifically, medications with higher AC potency are associated with elevated risk for ADRD.^13,14^ Exposure to higher-potency AC medications is also associated with elevated risk for falls^15,16^ and mortality.^17^ Autistic adults likely have higher and earlier exposure to medications with AC effects, attributable in part to high rates of co-occurring medical conditions for which such medications may be prescribed.^2,18,19^ Polypharmacy, including psychotropic polypharmacy, is common in autistic individuals,^20–27^ and prescription rates of psychotropic medications, which often have AC effects, increase with age.^26^

Despite evidence of poorer aging-related outcomes and high rates of polypharmacy, only two studies have examined AC medication use among autistic adults. These studies, which included 40+ year-old middle and older age autistic adults (*N*=415) without ID and 18-68-year-old autistic adults with ID aged (*N*=512), found that AC medication use was common and associated with self-reported^27^ and proxy-reported^28^ memory problems and declines in cognition/behavior, respectively.

No study has examined AC medication exposure and its associations with health-related outcomes among autistic adults in healthcare claims data, which allow for population-level estimates. Thus, it is unclear if AC medication exposure is more common among autistic relative to general population adults or if the patterning of associations between these medications and aging-related outcomes is the same or different among these populations.

The current study therefore takes a foundational first step, characterizing exposure to medications with AC effects and examining associations of AC exposure and potency with ADRD, falls, and mortality in a national US sample of autistic adults without intellectual disability (ID), autistic adults with ID, non-autistic adults with ID, and a comparison group with neither autism nor ID.

## Methods

This study used 2017-2019 Medicare and Medicaid data from 50 states and the District of Columbia, where sources were Medicare fee-for-service data and Medicaid data from Transformed Medicaid Statistical Information System (T-MSIS) Analytic Files (TAF). Datasets included demographic and enrollment information, as well as linked claims data covering diagnoses, healthcare utilization, and pharmacologic treatments. Individuals were linked across Medicare and Medicaid files using individual-level identifiers. The use of these data for research was approved via HIPAA waiver by the institutional review board of Drexel University (protocol #2501010962).

Study inclusionary criteria were the following: enrolled in Medicare, Medicaid, or dually enrolled in Medicare and Medicaid, with at least 12 months of enrollment in 2017-2019; aged ≥40 years; and meeting claims-based criteria using validated algorithms from the CMS Chronic Conditions Data Warehouse (CCW) for autism or ID, both autism and ID, or having no claims for these conditions (control).^4,5^ Additionally, Medicare-only beneficiaries whose Medicare coverage consisted exclusively of managed care during the study period were excluded because their claims were unavailable.

This study included not only adults ≥65 years but also those below the age of 65 (the more common age cutoff in studies of medications with AC effects), based on accumulating evidence that autistic people experience aging-related cognitive^3^ and physical health^5^ challenges earlier in the lifespan. As in prior research, persons with Down syndrome were excluded due to their genetic risk for ADRD.^29^

Autism and ID were ascertained using all available Medicare and Medicaid data, including data outside of the study period, because both are lifelong neurodevelopmental conditions. Autism and/or ID conditions required at least one inpatient claim or two non-inpatient claims, an approach aligned with prior claims data research.^30–32^ Individuals were identified as having ADRD or as having fallen if they had a single ADRD (using the Bynum-standard algorithm^33^) or fall-related claim, respectively. Mortality information was identified using indicators from the relevant Medicare and/or Medicaid databases. See Table S1 for ICD-10 codes and/or claims/indicators for these variables.

### Data Analysis

#### Medication Coding for AC Potency

Medications were determined based on Medicaid pharmacy claims and Medicare Part D prescription drug event claims. Exposure to medication(s) with AC effects was operationalized in two ways using the CRIDECO Anticholinergic Load Scale (CALS).^34^ First, AC medication exposure was dichotomized into no exposure to medication(s) with AC effects versus exposure to any medication with AC effects. Second, among those prescribed at least one medication with AC effects, AC potency was categorized according to the highest CALS score of any AC medication dispensed during the study period: low (AC=1), moderate (AC=2), or high (AC=3). See Supplementary Methods for details concerning medication coding, Table S2 for medication routes excluded from analyses, and Table S3 for a complete list of medications and their CALS potency scores.

#### Statistical Approach

Modified Poisson regressions with robust standard errors were conducted to obtain the prevalence ratios and 95% confidence intervals for each outcome: ADRD, falls, or mortality.^35^ Models included the four-level diagnostic group variable (autism-only, autism+ID, ID-only, control), each of the two methods of AC medication characterization (no exposure vs. any exposure; exposure of medications with low, moderate, high potency AC effects), and the interactions between diagnostic group and the corresponding AC medication characterizations served as the independent variables of interest.

Covariates were age group (40-44, 45-49, 50-54, 55-59, 60-64, 65-69, 70-79, and 80+ years), sex, race/ethnicity, enrolled months group (12-24, 25-36), enrollment type (Medicare, Medicaid, dually enrolled), and U.S. state of residence. Pairwise contrasts were used to compare AC exposure categories within each diagnostic group and diagnostic groups within the same AC exposure or potency category. Statistical significance was defined as a two-sided *p* value <.05.

#### Sensitivity Analyses

Because indications for some medications with AC effects (e.g., depression) could also confer risk for one of the outcomes of interest evaluated here (e.g., ADRD), the prevalence of these outcomes could be related to the indication of the medication rather than the medication itself.^36^ For instance, depression can be part of the dementia prodrome and may also confer risk for dementia,^37,38^ while antipsychotic medications may be prescribed for neuropsychiatric symptoms in dementia.^39^ Thus, we conducted modified Poisson regressions after excluding persons prescribed medications with AC effects categorized as antidepressants or antipsychotics to help moderate concerns regarding possible indication bias (see Table S4).

## Results

The final prevalence sample from the combined Medicare and Medicaid samples was comprised of 61,212,067 persons aged 40+ years (see Figure 1): autism-only *N*=48,839; autism+ID *N*=80,439; ID-only *N*=573,846; control *N=*60,508,943. Table 1 reports prevalence of exposure to medications with AC effects and outcomes (ADRD, falls, mortality), as well as demographic information within each group.

**Figure 1.**
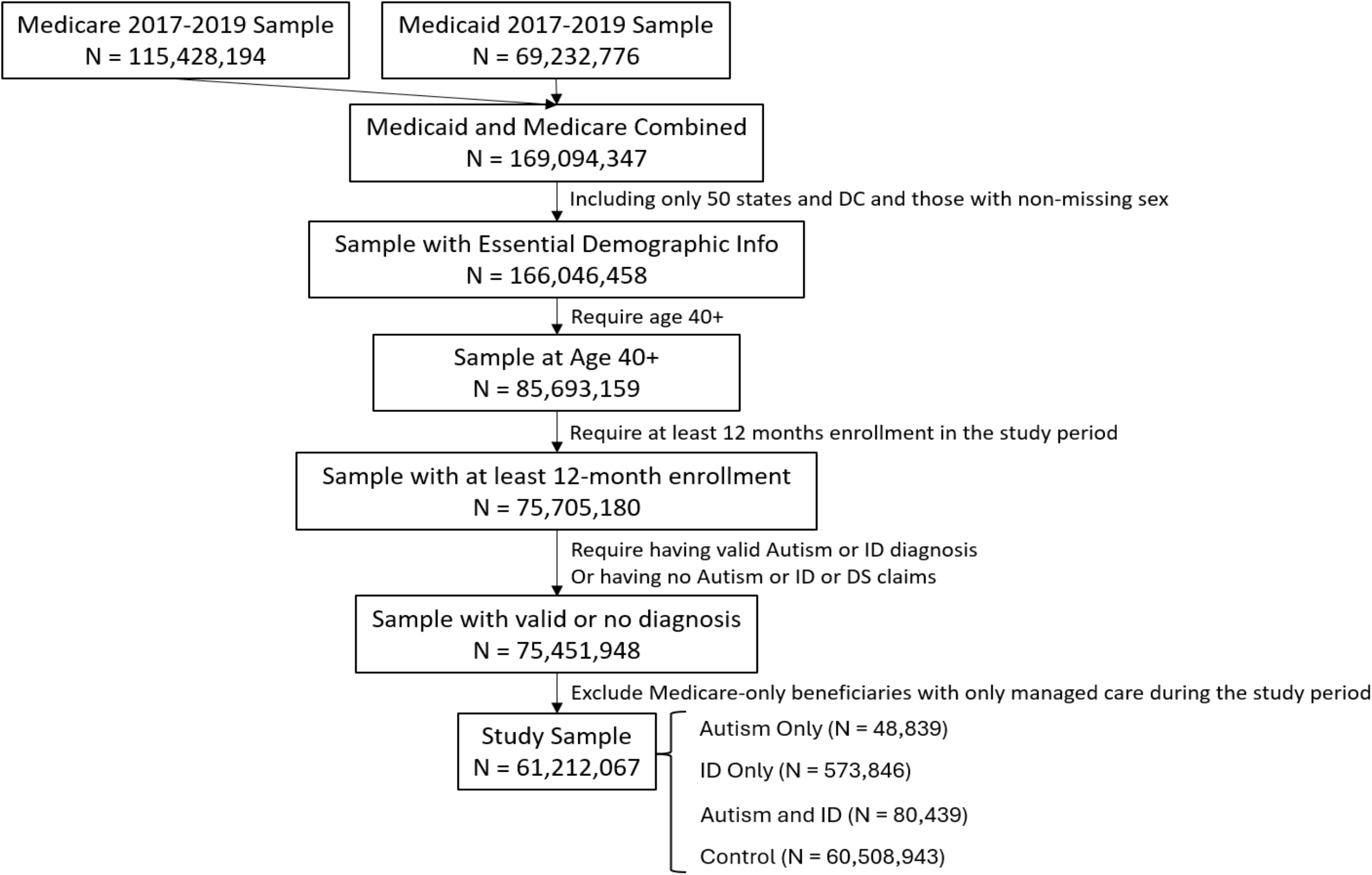
Flowchart indicating steps for inclusion/exclusion leading to the final sample for the current study.

**Figure 2.**
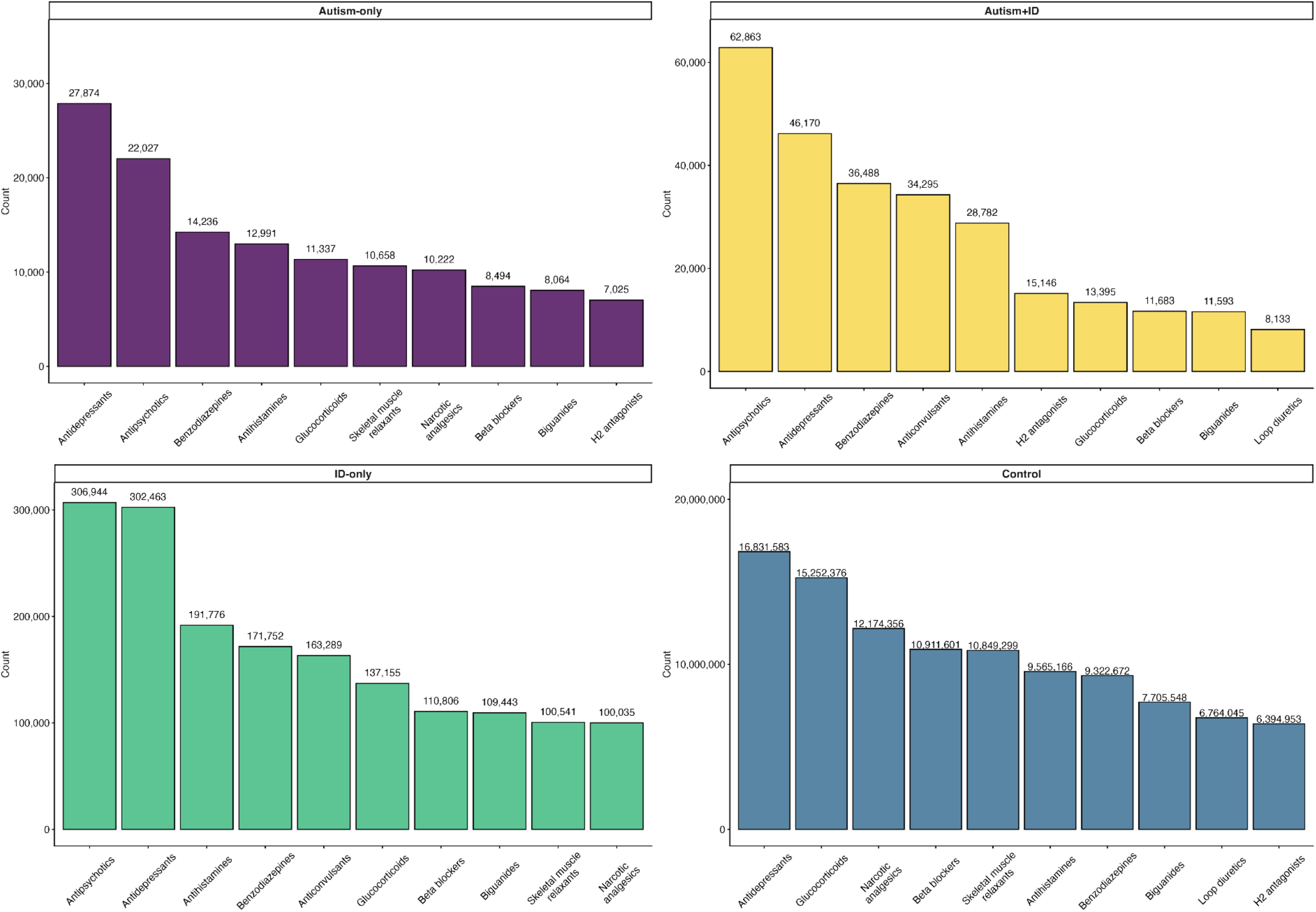
Visual depiction of the 10 most common medication classes with anticholinergic (AC) effects within each group (from top left to bottom right: autism-only, autism+intellectual disability (ID), ID-only, control), where AC classification was based on the CRIDECO Anticholinergic Load Scale (CALS)^34^. Note that counts represent the number of persons prescribed at least one medication in a given class with any AC potency and not the count of all AC medications within that class; thus, any given person may have been prescribed more than one medication in a relevant class (e.g., more than one antipsychotic), which is not reflected in the count. Note also that individuals may have been prescribed medications across two or more classes listed here (e.g., an individual may have been prescribed an antipsychotic, an antidepressant, and a benzodiazepine with AC effects).

**Table 1.** Characteristics of Participants in the Autism-only, Autism+ID, ID-only, and Control Groups.

|  | **Autism-only**<br>*N*=48,839 |  | **Autism+ID**<br>*N*=80,439 |  | **ID-only**<br>*N*=573,846 |  | **Control**<br>*N*=60,508,943 |  |
|---|---|---|---|---|---|---|---|---|
|  | **No AC Meds** | **AC Meds** | **No AC Meds** | **AC Meds** | **No AC Meds** | **AC Meds** | **No AC Meds** | **AC Meds** |
| ***n*, (%)** | 9,755 (19.97%) | 39,084 (80.02%) | 6,053 (7.52%) | 74,386 (92.48%) | 65,495 (11.41%) | 508,351 (88.59%) | 23,411,587 (38.69%) | 37,097,356 (61.30%) |
| **Age, years** |  |  |  |  |  |  |  |  |
| Mean (SD) | 55.91 (12.95) | 54.64 (12.22) | 50.46 (9.61) | 52.17 (9.69) | 54.58 (11.85) | 56.27 (11.27) | 65.37 (12.59) | 65.12 (12.99) |
| **Age Group, *n* (%)** |  |  |  |  |  |  |  |  |
| 40-44 | 2,471 (25.33) | 10,099 (25.84) | 2,089 (34.51) | 20,121 (27.05) | 16,220 (24.77) | 87,828 (17.28) | 2,081,112 (8.89) | 3,007,246 (8.11) |
| 45-49 | 1,316 (13.49) | 5,974 (15.29) | 1,161 (19.18) | 13,234 (17.79) | 9,841 (15.03) | 70,743 (13.92) | 1,313,388 (5.61) | 2,486,406 (6.70) |
| 50-54 | 1,288 (13.20) | 5,619 (14.38) | 1,002 (16.55) | 13,029 (17.52) | 9,864 (15.06) | 82,050 (16.14) | 1,231,754 (5.26) | 2,933,832 (7.91) |
| 55-59 | 964 (9.88) | 5,047 (12.91) | 713 (11.78) | 11,131 (14.96) | 9,097 (13.89) | 83,497 (16.43) | 1,284,103 (5.48) | 3,272,351 (8.82) |
| 60-64 | 1,025 (10.51) | 4,147 (10.61) | 511 (8.44) | 8,177 (10.99) | 7,263 (11.09) | 67,910 (13.36) | 3,756,502 (16.05) | 4,852,051 (13.08) |
| 65-69 | 1,113 (11.41) | 3,164 (8.10) | 318 (5.25) | 4,725 (6.35) | 5,467 (8.35) | 50,018 (9.84) | 5,773,917 (24.66) | 6,782,678 (18.28) |
| 70-79 | 1,103 (11.31) | 3,424 (8.76) | 207 (3.42) | 3,402 (4.57) | 5,383 (8.22) | 49,605 (9.76) | 5,225,278 (22.32) | 8,697,772 (23.45) |
| 80+ | 475 (4.87) | 1,610 (4.12) | 52 (0.86) | 567 (0.76) | 2,360 (3.60) | 16,700 (3.29) | 2,745,533 (11.73) | 5,065,020 (13.65) |
| **Sex, *n* (%)** |  |  |  |  |  |  |  |  |
| Female | 2,572 (26.37) | 14,637 (36.76) | 1,623 (26.81) | 24,618 (33.09) | 26,257 (40.09) | 245,087 (48.21) | 11,154,046 (47.64) | 21,984,090 (59.26) |
| Male | 7,183 (73.63) | 24,717 (63.24) | 4,430 (73.19) | 49,768 (66.91) | 39,238 (59.91) | 263,264 (51.79) | 12,257,541 (52.36) | 15,113,266 (40.74) |
| **Race/Ethnicity, *n* (%)** |  |  |  |  |  |  |  |  |
| Asian/Pacific Islander | 198 (2.03) | 842 (2.15) | 139 (2.30) | 1,308 (1.76) | 1,855 (2.83) | 9,887 (1.94) | 1,063,668 (4.54) | 1,741,428 (4.69) |
| Black | 1,060 (10.87) | 3,829 (9.80) | 1,415 (23.32) | 12,744 (17.13) | 15,374 (23.47) | 103,320 (20.32) | 2,576,968 (11.01) | 4,865,827 (13.12) |
| Hispanic/Latino | 436 (4.47) | 2,312 (5.92) | 413 (6.82) | 4,541 (6.10) | 5,801 (8.86) | 40,150 (7.90) | 2,700,173 (11.53) | 4,267,763 (11.50) |
| Native American /Alaska Native | 60 (0.62) | 264 (0.68) | 34 (0.56) | 441 (0.59) | 747 (1.14) | 4,547 (0.89) | 179,237 (0.77) | 257,306 (0.69) |
| Multiracial/Other | 57 (0.58) | 129 (0.33) | *15 (0.25)* | 106 (0.14) | 185 (0.28) | 809 (0.16) | 180,522 (0.77) | 155,569 (0.42) |
| Missing | 319 (3.27) | 932 (2.38) | 67 (1.11) | 691 (0.93) | 936 (1.43) | 6,594 (1.30) | 1,267,670 (5.41) | 1,313,908 (3.54) |
| White | 7,625 (78.17) | 30,776 (78.84) | 3,970 (65.59) | 54,555 (73.34) | 40,597 (61.98) | 343,044 (67.48) | 15,443,349 (65.96) | 24,495,555 (66.03) |
| **Enrolled months** |  |  |  |  |  |  |  |  |
| Mean (SD) | 32.48 (6.79) | 33.27 (6.01) | 33.35 (6.09) | 34.11 (5.18) | 33.43 (6.00) | 34.02 (5.26) | 30.96 (7.91) | 32.82 (6.39) |
| **Enrolled months group** |  |  |  |  |  |  |  |  |
| 12-24 | 1,610 (16.50) | 4,796 (12.27) | 772 (12.75) | 6,524 (8.77) | 8,137 (12.42) | 46,681 (9.18) | 5,458,049 (23.31) | 5,275,735 (14.22) |
| 25-36 | 8,145 (83.50) | 34,288 (87.73) | 5,281 (87.25) | 67,862 (91.23) | 57,358 (87.58) | 461,670 (90.82) | 17,953,538 (76.69) | 31,821,621 (85.78) |
| **Enrollment Type, *n* (%)** |  |  |  |  |  |  |  |  |
| Medicaid Only | 2,018 (20.69) | 10,451 (26.74) | 914 (15.10) | 9,740 (13.09) | 11,340 (17.31) | 92,180 (18.13) | 5,027,385 (21.47) | 9,004,552 (24.27) |
| Medicare Only | 4,700 (48.18) | 6,066 (15.52) | 912 (15.07) | 1,218 (1.64) | 15,604 (23.82) | 19,359 (3.81) | 16,875,741(72.08) | 16,811,030 (45.32) |
| Dually Enrolled | 3,037 (31.13) | 22,567 (57.74) | 4,227 (69.83) | 63,428 (85.27) | 38,551 (58.86) | 396,812 (78.06) | 1,508,461 (6.44) | 11,281,774 (30.41) |
| **Region, *n* (%)** |  |  |  |  |  |  |  |  |
| Midwest | 2,023 (20.74) | 9,351 (23.93) | 1,212 (20.02) | 18,307 (24.61) | 14,864 (22.69) | 134,327 (26.42) | 4,421,900 (18.89) | 8,116,085 (21.88) |
| Northeast | 2,123 (21.76) | 9,392 (24.03) | 1,676 (27.69) | 21,481 (28.88) | 14,307 (21.84) | 111,094 (21.85) | 4,418,282 (18.87) | 7,795,891 (21.01) |
| South | 2,986 (30.61) | 10,218 (26.14) | 2,146 (35.45) | 22,314 (30.00) | 24,150 (36.87) | 182,719 (35.94) | 8,238,427 (35.19) | 13,015,859 (35.09) |
| West | 2,623 (26.89) | 10,123 (25.90) | 1,019 (16.83) | 12,284 (16.51) | 12,174 (18.59) | 80,211 (15.78) | 6,332,978 (27.05) | 8,169,521 (22.02) |
| **Conditions, *n* (%)** |  |  |  |  |  |  |  |  |
| ADRD | 809 (8.29) | 5,577 (14.27) | 278 (4.59) | 10,969 (14.75) | 5,289 (8.08) | 105,495 (20.75) | 1,258,382 (5.38) | 4,127,775 (11.13) |
| Fall | 813 (8.33) | 5,852 (14.97) | 462 (7.63) | 16,663 (22.40) | 6,168 (9.42) | 112,713 (22.17) | 1,587,748 (6.78) | 4,658,409 (12.56) |
| Mortality | 379 (3.89) | 2,144 (5.49) | 140 (2.31) | 3,591 (4.83) | 2,746 (4.19) | 41,666 (8.20) | 1,084,893 (4.63) | 2,795,714 (7.54) |

Abbreviations. AC=anticholinergic; ADRD=Alzheimer disease and related dementias; Dually Enrolled=dually enrolled in Medicare and Medicaid; ID=intellectual disability.

### Characterization of Medications with AC Effects

Individuals in the autism-only, autism+ID, and ID-only groups were all significantly more likely to be prescribed at least one medication with AC effects compared to individuals in the control group (*p*s<.05). The patterning of medication exposure across groups, where each pairwise comparison was significant, was as follows: autism+ID (92.5%) > ID-only (88.6%) > autism-only (80.0%) > non-autism/non-ID controls (61.3%).

The highest frequency categories of medications with AC effects for each group are visualized in Figure 1. Across all groups, antidepressants were among the most frequent. Antipsychotic medications were the most common in the autism+ID and ID-only groups and second most common (after antidepressants) within the autism-only group.

### Associations of AC Medication Exposure (any vs. none), Group (autism, autism+ID, ID, control), and their Interaction with ADRD, Falls, and Mortality Prevalence

Relative to the control group not prescribed any AC medication, across the three clinical groups (autism-only, autism+ID, ID-only), ADRD, falls and mortality were all more frequent for those prescribed at least one medication with AC effects (Table 2). Within each diagnostic group, persons prescribed any medication with AC effects compared to those prescribed no such medication were more likely to have an ADRD or fall-related claim, and more likely to experience death (Table 3A). The interaction between diagnostic group and AC medication exposure was statistically significant for all three outcomes (Table 2) and follow-up pairwise comparisons among AC-exposed individuals showed that the autism-only, autism+ID, and ID-only groups each had higher adjusted prevalences of ADRD, falls, and mortality than the control group (Table 3B).

**Table 2.**
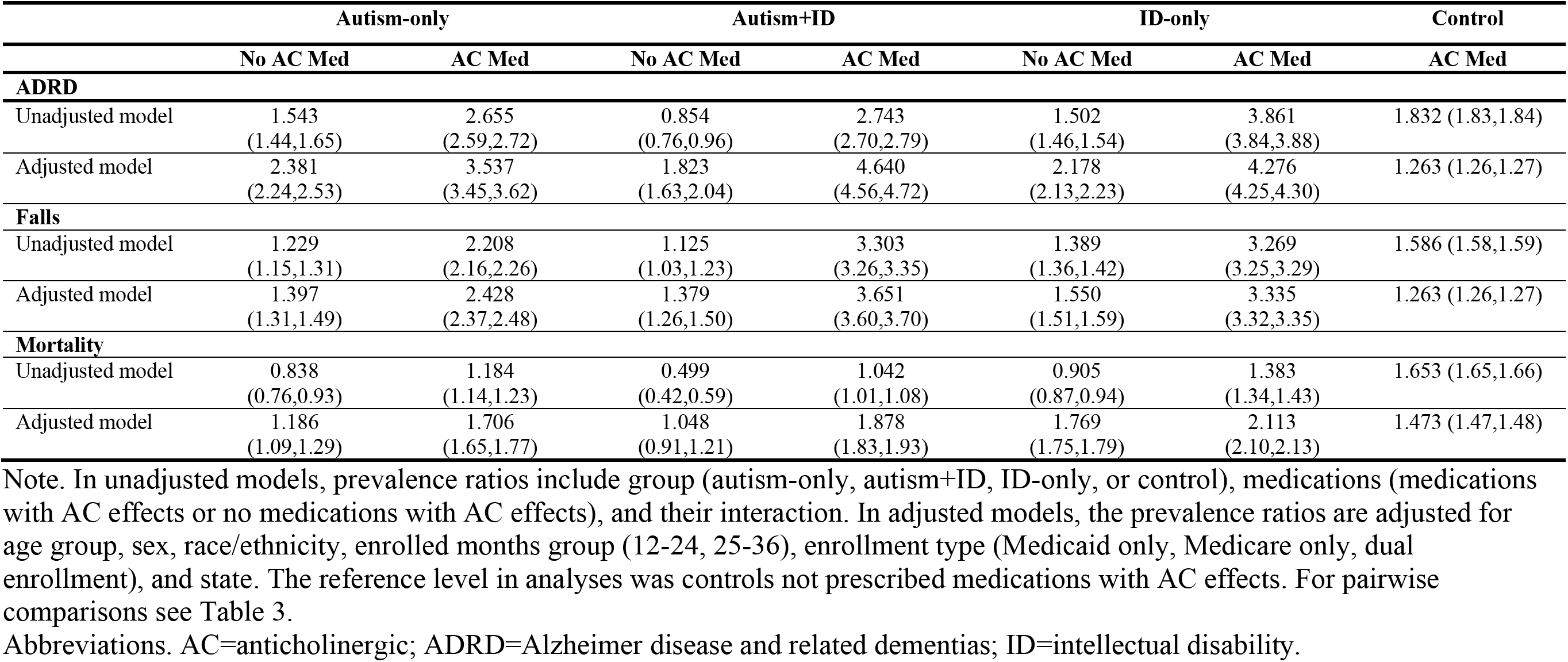
Prevalence Ratios and 95% Confidence Intervals of Alzheimer Disease and Related Dementias (ADRD), Falls, and Mortality for Persons not Prescribed Medications with Anticholinergic (AC) Effects and Persons Prescribed at Least One Medication with AC Effects for 2017-2019 Medicaid and/or Medicare Enrollees aged ≥40 Years in the Four Groups.

|  | Autism-only |  | Autism+ID |  | ID-only |  | Control |
|---|---|---|---|---|---|---|---|
|  | **No AC Med** | **AC Med** | **No AC Med** | **AC Med** | **No AC Med** | **AC Med** | **AC Med** |
| **ADRD** |  |  |  |  |  |  |  |
| Unadjusted model | 1.543 (1.44,1.65) | 2.655 (2.59,2.72) | 0.854 (0.76,0.96) | 2.743 (2.70,2.79) | 1.502 (1.46,1.54) | 3.861 (3.84,3.88) | 1.832 (1.83,1.84) |
| Adjusted model | 2.381 (2.24,2.53) | 3.537 (3.45,3.62) | 1.823 (1.63,2.04) | 4.640 (4.56,4.72) | 2.178 (2.13,2.23) | 4.276 (4.25,4.30) | 1.263 (1.26,1.27) |
| **Falls** |  |  |  |  |  |  |  |
| Unadjusted model | 1.229 (1.15,1.31) | 2.208 (2.16,2.26) | 1.125 (1.03,1.23) | 3.303 (3.26,3.35) | 1.389 (1.36,1.42) | 3.269 (3.25,3.29) | 1.586 (1.58,1.59) |
| Adjusted model | 1.397 (1.31,1.49) | 2.428 (2.37,2.48) | 1.379 (1.26,1.50) | 3.651 (3.60,3.70) | 1.550 (1.51,1.59) | 3.335 (3.32,3.35) | 1.263 (1.26,1.27) |
| **Mortality** |  |  |  |  |  |  |  |
| Unadjusted model | 0.838 (0.76,0.93) | 1.184 (1.14,1.23) | 0.499 (0.42,0.59) | 1.042 (1.01,1.08) | 0.905 (0.87,0.94) | 1.383 (1.34,1.43) | 1.653 (1.65,1.66) |
| Adjusted model | 1.186 (1.09,1.29) | 1.706 (1.65,1.77) | 1.048 (0.91,1.21) | 1.878 (1.83,1.93) | 1.769 (1.75,1.79) | 2.113 (2.10,2.13) | 1.473 (1.47,1.48) |

Note. In unadjusted models, prevalence ratios include group (autism-only, autism+ID, ID-only, or control), medications (medications with AC effects or no medications with AC effects), and their interaction. In adjusted models, the prevalence ratios are adjusted for age group, sex, race/ethnicity, enrolled months group (12-24, 25-36), enrollment type (Medicaid only, Medicare only, dual enrollment), and state. The reference level in analyses was controls not prescribed medications with AC effects. For pairwise comparisons see Table 3.
Abbreviations. AC=anticholinergic; ADRD=Alzheimer disease and related dementias; ID=intellectual disability.

**Table 3.**
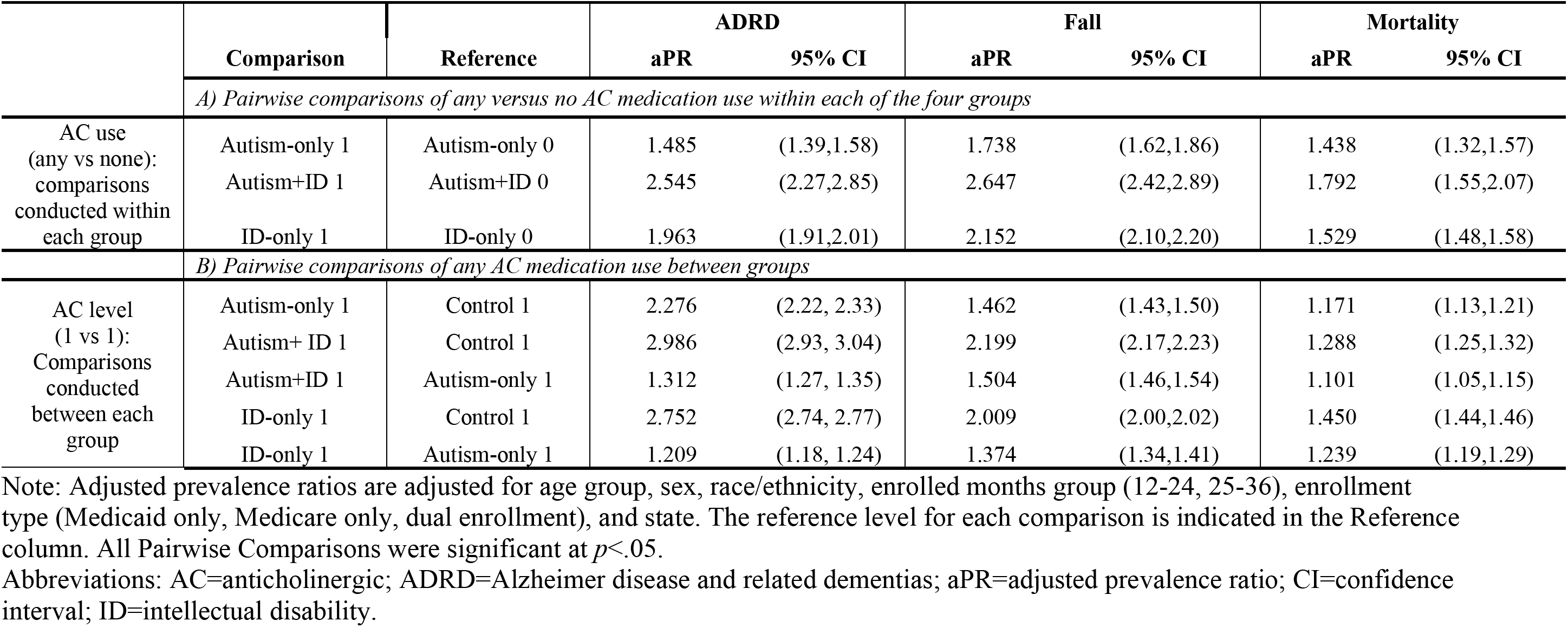
Adjusted Prevalence Ratios (aPR) and 95% Confidence Intervals (CI) of Alzheimer Disease and Related Dementias (ADRD), Falls, and Mortality for Pairwise Comparisons for A) None versus any Anticholinergic (AC) Medication Exposure within each Group (e.g., Autism AC=1 versus Autism AC=0); and B) any AC Medication Use by Group (e.g., Autism AC=1 versus Control AC=1).

|  |  |  | ADRD |  | Fall |  | Mortality |  |
|---|---|---|---|---|---|---|---|---|
|  | Comparison | Reference | aPR | 95% CI | aPR | 95% CI | aPR | 95% CI |
|  | *A) Pairwise comparisons of any versus no AC medication use within each of the four groups* |  |  |  |  |  |  |  |
| AC use<br>(any vs none):<br>comparisons conducted within each group | Autism-only 1 | Autism-only 0 | 1.485 | (1.39,1.58) | 1.738 | (1.62,1.86) | 1.438 | (1.32,1.57) |
|  | Autism+ID 1 | Autism+ID 0 | 2.545 | (2.27,2.85) | 2.647 | (2.42,2.89) | 1.792 | (1.55,2.07) |
|  | ID-only 1 | ID-only 0 | 1.963 | (1.91,2.01) | 2.152 | (2.10,2.20) | 1.529 | (1.48,1.58) |
|  | *B) Pairwise comparisons of any AC medication use between groups* |  |  |  |  |  |  |  |
| AC level<br>(1 vs 1):<br>Comparisons conducted between each group | Autism-only 1 | Control 1 | 2.276 | (2.22, 2.33) | 1.462 | (1.43,1.50) | 1.171 | (1.13,1.21) |
|  | Autism+ ID 1 | Control 1 | 2.986 | (2.93, 3.04) | 2.199 | (2.17,2.23) | 1.288 | (1.25,1.32) |
|  | Autism+ID 1 | Autism-only 1 | 1.312 | (1.27, 1.35) | 1.504 | (1.46,1.54) | 1.101 | (1.05,1.15) |
|  | ID-only 1 | Control 1 | 2.752 | (2.74, 2.77) | 2.009 | (2.00,2.02) | 1.450 | (1.44,1.46) |
|  | ID-only 1 | Autism-only 1 | 1.209 | (1.18, 1.24) | 1.374 | (1.34,1.41) | 1.239 | (1.19,1.29) |

Note: Adjusted prevalence ratios are adjusted for age group, sex, race/ethnicity, enrolled months group (12-24, 25-36), enrollment type (Medicaid only, Medicare only, dual enrollment), and state. The reference level for each comparison is indicated in the Reference column. All Pairwise Comparisons were significant at *p*<.05.
Abbreviations: AC=anticholinergic; ADRD=Alzheimer disease and related dementias; aPR=adjusted prevalence ratio; CI=confidence interval; ID=intellectual disability.

### Associations of AC Potency Level (1, 2, 3) with ADRD, Falls, and Mortality

Relative to the control group prescribed a medication with an AC potency of 1, all three clinical groups of autism-only, autism+ID, and ID-only, had higher adjusted prevalences of ADRD, falls, or mortality (Table 4). Within each of the autism-only, autism+ID, and ID-only groups, higher potency AC medications were significantly associated with a stepwise increase in the prevalence of ADRD and falls (i.e., AC=3 > AC=2 > AC=1) (Table 5A). Mortality prevalence also generally increased with AC potency. The interaction between AC medication potency level and group was statistically significant for ADRD, falls, and mortality (Table 4). Pairwise follow-up comparisons indicated that relative to control persons prescribed the same level of AC medication potency, each clinical group broadly had a higher prevalence of ADRD, falls, and mortality (Table 5B).

**Table 4.**
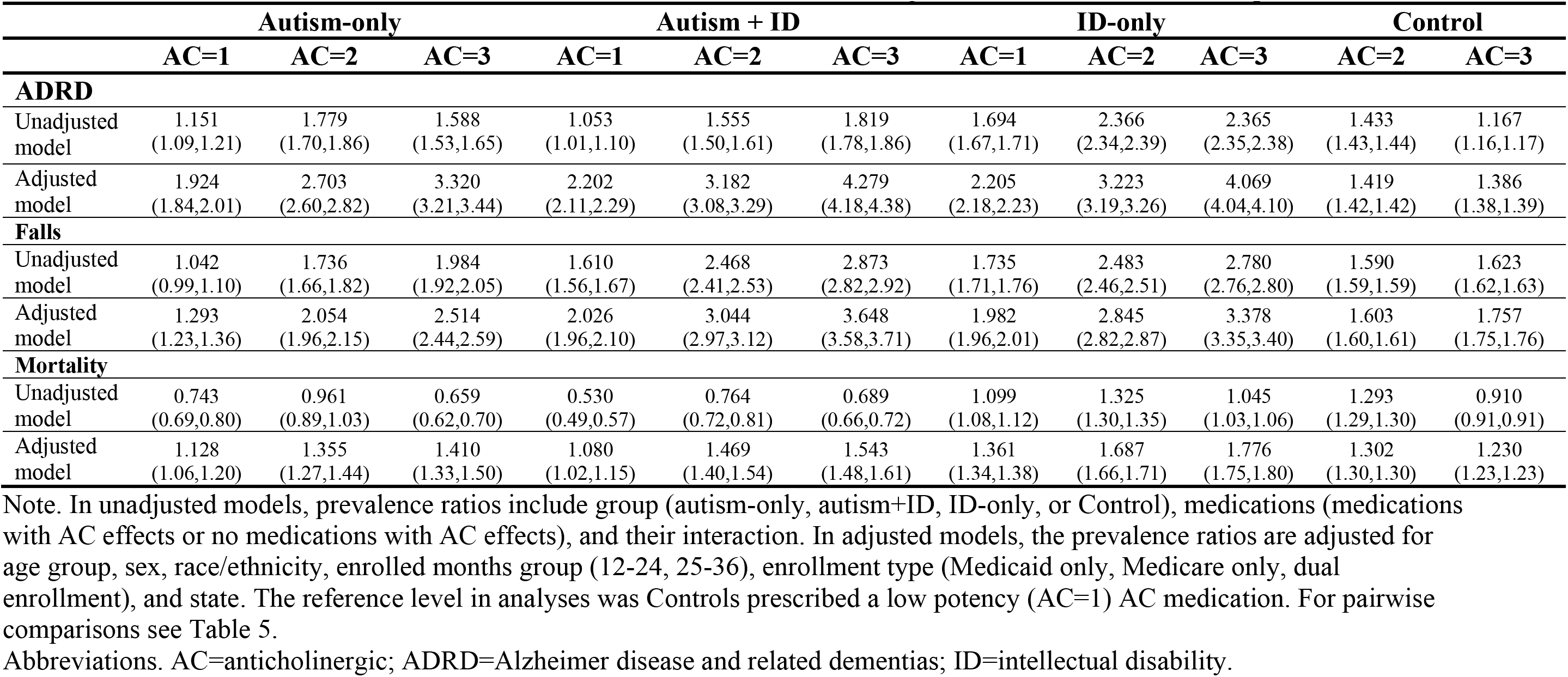
Prevalence Ratios and 95% Confidence Intervals of Alzheimer Disease and Related Dementias (ADRD), Falls, and Mortality for Anticholinergic (AC) Potency Levels of Low, Medium, and High (AC=1, AC=2, AC=3) for Persons Prescribed at Least One Medication with AC Effects for 2017-2019 Medicaid and/or Medicare Enrollees Aged ≥40 Years in the Four Groups.

|  | Autism-only |  |  | Autism + ID |  |  | ID-only |  |  | Control |  |
|---|---|---|---|---|---|---|---|---|---|---|---|
|  | AC=1 | AC=2 | AC=3 | AC=1 | AC=2 | AC=3 | AC=1 | AC=2 | AC=3 | AC=2 | AC=3 |
| **ADRD** |  |  |  |  |  |  |  |  |  |  |  |
| Unadjusted model | 1.151<br>(1.09,1.21) | 1.779<br>(1.70,1.86) | 1.588<br>(1.53,1.65) | 1.053<br>(1.01,1.10) | 1.555<br>(1.50,1.61) | 1.819<br>(1.78,1.86) | 1.694<br>(1.67,1.71) | 2.366<br>(2.34,2.39) | 2.365<br>(2.35,2.38) | 1.433<br>(1.43,1.44) | 1.167<br>(1.16,1.17) |
| Adjusted model | 1.924<br>(1.84,2.01) | 2.703<br>(2.60,2.82) | 3.320<br>(3.21,3.44) | 2.202<br>(2.11,2.29) | 3.182<br>(3.08,3.29) | 4.279<br>(4.18,4.38) | 2.205<br>(2.18,2.23) | 3.223<br>(3.19,3.26) | 4.069<br>(4.04,4.10) | 1.419<br>(1.42,1.42) | 1.386<br>(1.38,1.39) |
| **Falls** |  |  |  |  |  |  |  |  |  |  |  |
| Unadjusted model | 1.042<br>(0.99,1.10) | 1.736<br>(1.66,1.82) | 1.984<br>(1.92,2.05) | 1.610<br>(1.56,1.67) | 2.468<br>(2.41,2.53) | 2.873<br>(2.82,2.92) | 1.735<br>(1.71,1.76) | 2.483<br>(2.46,2.51) | 2.780<br>(2.76,2.80) | 1.590<br>(1.59,1.59) | 1.623<br>(1.62,1.63) |
| Adjusted model | 1.293<br>(1.23,1.36) | 2.054<br>(1.96,2.15) | 2.514<br>(2.44,2.59) | 2.026<br>(1.96,2.10) | 3.044<br>(2.97,3.12) | 3.648<br>(3.58,3.71) | 1.982<br>(1.96,2.01) | 2.845<br>(2.82,2.87) | 3.378<br>(3.35,3.40) | 1.603<br>(1.60,1.61) | 1.757<br>(1.75,1.76) |
| **Mortality** |  |  |  |  |  |  |  |  |  |  |  |
| Unadjusted model | 0.743<br>(0.69,0.80) | 0.961<br>(0.89,1.03) | 0.659<br>(0.62,0.70) | 0.530<br>(0.49,0.57) | 0.764<br>(0.72,0.81) | 0.689<br>(0.66,0.72) | 1.099<br>(1.08,1.12) | 1.325<br>(1.30,1.35) | 1.045<br>(1.03,1.06) | 1.293<br>(1.29,1.30) | 0.910<br>(0.91,0.91) |
| Adjusted model | 1.128<br>(1.06,1.20) | 1.355<br>(1.27,1.44) | 1.410<br>(1.33,1.50) | 1.080<br>(1.02,1.15) | 1.469<br>(1.40,1.54) | 1.543<br>(1.48,1.61) | 1.361<br>(1.34,1.38) | 1.687<br>(1.66,1.71) | 1.776<br>(1.75,1.80) | 1.302<br>(1.30,1.30) | 1.230<br>(1.23,1.23) |

Note. In unadjusted models, prevalence ratios include group (autism-only, autism+ID, ID-only, or Control), medications (medications with AC effects or no medications with AC effects), and their interaction. In adjusted models, the prevalence ratios are adjusted for age group, sex, race/ethnicity, enrolled months group (12-24, 25-36), enrollment type (Medicaid only, Medicare only, dual enrollment), and state. The reference level in analyses was Controls prescribed a low potency (AC=1) AC medication. For pairwise comparisons see Table 5.
Abbreviations. AC=anticholinergic; ADRD=Alzheimer disease and related dementias; ID=intellectual disability.

**Table 5.** Adjusted Prevalence Ratios (aPR) and 95% Confidence Intervals (CI) of Alzheimer Disease and Related Dementias (ADRD), Falls, and Mortality for Pairwise Comparisons for A) Anticholinergic (AC) Medication Exposure Potency (AC=1, AC=2, AC=3) within each Group (e.g., Autism AC=1 vs. Autism AC=2); and B) Medication Exposure Potency between Groups (e.g., Autism AC=3 vs. Control AC=3).

|  |  |  | ADRD |  | Fall |  | Mortality |  |
|---|---|---|---|---|---|---|---|---|
|  | Comparison | Reference | aPR | 95% CI | aPR | 95% CI | aPR | 95% CI |
|  | *A) Pairwise comparisons for AC medication exposure potency (AC=1, AC=2, AC=3) within each of the four groups* |  |  |  |  |  |  |  |
| AC potency (1, 2, 3): comparisons conducted within each group | Control 2 | Control 3 | 1.024 | (1.02,1.03) | 0.912 | (0.91,0.91) | 1.058 | (1.06,1.06) |
|  | Autism-only 1 | Autism-only 2 | 0.712 | (0.67,0.76) | 0.629 | (0.59,0.67) | 0.833 | (0.76,0.91) |
|  | Autism-only 2 | Autism-only 3 | 0.814 | (0.77,0.86) | 0.817 | (0.77,0.86) | 0.961 | (0.88,1.05) |
|  | Autism+ID 1 | Autism+ID 2 | 0.692 | (0.66,0.73) | 0.666 | (0.64,0.69) | 0.735 | (0.68,0.79) |
|  | Autism+ID 2 | Autism+ID 3 | 0.744 | (0.71,0.77) | 0.835 | (0.81,0.86) | 0.953 | (0.89,1.01) |
|  | ID-only 1 | ID-only 2 | 0.684 | (0.67,0.69) | 0.697 | (0.69,0.71) | 0.807 | (0.79,0.82) |
|  | ID-only 2 | ID-only 3 | 0.792 | (0.78,0.80) | 0.842 | (0.83,0.85) | 0.950 | (0.93,0.97) |
|  | *B) Pairwise comparisons AC medication exposure potency (AC=1, AC=2, AC=3) between each of the four groups* |  |  |  |  |  |  |  |
| AC potency (1,2,3): Comparisons conducted between each group | Autism-only 2 | Control 2 | 1.905 | (1.83,1.98) | 1.281 | (1.23,1.34) | 1.041 | (0.98,1.11) |
|  | Autism-only 3 | Control 3 | 2.395 | (2.31,2.48) | 1.431 | (1.39,1.48) | 1.146 | (1.08,1.22) |
|  | Autism+ID 2 | Control 2 | 2.242 | (2.17,2.32) | 1.899 | (1.85,1.95) | 1.129 | (1.08,1.19) |
|  | Autism+ID 3 | Control 3 | 3.087 | (3.01,3.16) | 2.076 | (2.04,2.11) | 1.254 | (1.20,1.31) |
|  | ID-only 2 | Control 2 | 2.272 | (2.25,2.29) | 1.775 | (1.76,1.79) | 1.296 | (1.28,1.32) |
|  | ID-only 3 | Control 3 | 2.935 | (2.91,2.96) | 1.922 | (1.91,1.94) | 1.444 | (1.43,1.46) |
|  | Autism-only 1 | Autism+ID 1 | 0.874 | (0.82,0.93) | 0.638 | (0.60,0.68) | 1.045 | (0.96,1.14) |
|  | Autism-only 2 | Autism+ID 2 | 0.850 | (0.81,0.90) | 0.675 | (0.64,0.71) | 0.922 | (0.85,1.00) |
|  | Autism-only 3 | Autism+ID 3 | 0.776 | (0.74,0.81) | 0.689 | (0.67,0.71) | 0.914 | (0.85,0.98) |
|  | Autism+ID 1 | ID-only 1 | 0.998 | (0.96,1.04) | 1.022 | (0.99,1.06) | 0.794 | (0.75,0.84) |
|  | Autism+ID 2 | ID-only 2 | 0.987 | (0.95,1.02) | 1.070 | (1.04,1.10) | 0.871 | (0.83,0.92) |
|  | Autism+ID 3 | ID-only 3 | 1.052 | (1.03,1.08) | 1.080 | (1.06,1.10) | 0.869 | (0.83,0.91) |

Note: Adjusted prevalence ratios are adjusted for age group, sex, race/ethnicity, enrolled months group (12-24, 25-36), enrollment type (Medicaid only, Medicare only, dual enrollment), and state. The reference level for each comparison is indicated in the Reference column. All pairwise comparisons were significant at *p*<.05.
Abbreviations: AC=anticholinergic; ADRD=Alzheimer disease and related dementias; aPR=adjusted prevalence ratio; CI=confidence interval; ID=intellectual disability.

Sensitivity analyses removing individuals prescribed antidepressant and/or antipsychotic medication(s) with AC effects across the four groups (see Table S4 for subsample demographic characteristics) indicated findings between the clinical groups and the control group generally remained, although some exposure-outcome estimates were attenuated and the stepwise potency pattern was not consistently retained, particularly for mortality (see Supplementary Materials and Tables S5 and S6).

## Discussion

Using US nationwide healthcare claims data, this study provides a foundational first step in characterizing exposure to medications with AC effects and establishing associations between these medications and negative aging-related outcomes for which autistic people with and without ID and people with ID (without autism) are at increased risk (i.e., ADRD, falls, mortality). Overall, the current study found elevated rates of exposure to medications with AC effects in the clinical groups compared to the control group and links between AC exposure (bivariate)/AC potency (ordinal, stepwise) and prevalence ADRD, falls, and mortality, which were accentuated in the three clinical groups relative to the control group for several outcomes.

### Exposure to Medications with AC Effects

In the current sample of controls enrolled in Medicare, Medicaid or dually enrolled in Medicare and Medicaid, prevalence of exposure to AC medications was 61.3%. In the two completed studies that examined use of medications with AC effects among autistic adults with and without ID, rates of caregiver-reported and self-reported use of these medications was 66.4%^28^ and 62.9%,^27^ respectively. These rates are lower than those identified in the current claims-based data for autistic persons (autism-only=80.0%; autism+ID=92.5%), as well as for non-autistic persons with ID (88.6%). A potential explanation for higher rates in the current study relative to these prior studies may be related to the more complex medical needs of Medicare and/or Medicaid enrollees. For instance, persons who are <65 years and enrolled in Medicare (or dually enrolled in Medicare and Medicaid) must have a qualifying disability. It is possible that these persons experience greater levels of difficulty in functioning due to one or more conditions (including autism and/or ID), and so they may be prescribed more medications with AC effects than persons <65 enrolled in private health insurance.

This study demonstrates that the prevalence of exposure to medications with AC effects was higher for the clinical groups relative to the control group, where persons in the autism+ID group experienced the greatest exposure to such medications, and those in the control group the lowest. Results for the ID-only group are consistent with a retrospective cohort study of Scottish persons aged 17-94 years with an ID (*n*=4,305) and controls (*n*=12,915) matched for demographic characteristics, which indicated that persons with ID were significantly more likely to be prescribed medications with AC effects relative to persons without ID (OR=1.49 [1.38– 1.59]).^40^

In terms of classes of AC medications, as has been reported for 65+ year old adults in the general aging population,^41^ the current study found that for the three clinical groups and the control group antidepressants were among the most frequently prescribed class of medications with AC effects.^41^ Differing from the current study’s control sample and prior studies in the general aging population, but broadly consistent with reports of high rates of psychotropic medication use in autistic and ID samples^24–26,42^ and aligned with the prior two studies of AC medication use among autistic adults,^27,28^ antipsychotics^24–26^ were common for the autism-only, autism+ID, and ID-only groups. Concerns have been raised regarding the prescribing of psychotropic medications broadly and antipsychotic medications specifically among persons with ID.^43^ The current study’s findings underscore the need to better understand the “off-label” prescribing of antipsychotic medications in autistic persons and persons with ID, as we currently lack a firm knowledgebase regarding the risks and benefits of antipsychotic medications in these populations.

### Exposure to Medications with AC Effects (vs. No Exposure) and ADRD, Falls, and Mortality Prevalence

Across all groups, exposure to medications with AC effects compared to no such exposure was associated with greater frequency of ADRD, falls, and mortality. This finding is consistent with literature demonstrating associations between AC medication exposure and ADRD,^44,45^ falls,^15,16^ and mortality^17^ in the general aging population, and extends these links to autism and ID. For example, the findings of associations with AC medications and falls in persons with ID is concordant with a study of 487 Irish adults (aged 40+ years) with ID, which found AC medications were linked to higher odds of falls (1.86 [1.03–3.38], *p*=.04).^46^ However, this prior study in persons with ID identified lower odds of dementia/Alzheimer’s Disease (0.21 [0.07–0.64], *p*=.006) with AC medication use, though the authors noted limitations of their study, including the small number of persons reporting dementia/Alzheimer’s (*n*=29).^46^

In the current study, among individuals prescribed at least one AC medication, pairwise comparisons indicated that the autism-only, autism+ID, and ID-only groups each had higher adjusted prevalences of ADRD, falls, and mortality than the control group.

### AC Potency and ADRD, Falls, and Mortality Prevalence

In addition to findings of associations between AC medication exposure and aging-related outcomes, results suggested a stepwise pattern, wherein increasing levels of medication potency (AC=1<2<3) are broadly associated with increases in the burden of ADRD, falls, and mortality within the four groups. This finding is aligned with and extends (to autism and ID groups) the literature suggesting that medications with greater AC potency are associated with corresponding greater risks for dementia, falls/fractures and mortality in the general population.^47–49^ Additionally, pairwise comparisons indicated that at each level of AC potency, the autism-only, autism+ID, and ID-only groups broadly demonstrated greater prevalence ratios for ADRD, falls, and mortality relative to controls. For example, among those prescribed at least one medication with an AC potency of 3, prevalence of ADRD was 2.40, 3.09, and 2.94 times that in the control group for those in the autism-only, autism+ID, and ID-only groups, respectively.

A better understanding of potential reasons that people with neurodevelopmental conditions appear to show higher prevalence of ADRD, falls, and mortality associated with AC medication use relative to the general population is needed. It is possible that medications with AC effects could have differential effects among persons with neurodevelopmental conditions relative to persons without these conditions, in part due to vulnerability of underlying neurological systems in the former.

In the general aging population reasons for associations between AC medication exposure and fall risk are not fully clear; however, negative impacts of AC medications on balance and gait have been identified as a potential mechanism.^50^ Autistic persons and persons with ID may demonstrate differences in motor development; thus, impacts of AC medications on motor functioning may have particularly pronounced effects in persons with developmental motor differences, whether early-onset^51^ or emerging later in development.^52^

Similarly, persons with neurodevelopmental conditions such as autism and/or ID can demonstrate differences in cognition, including in executive functioning.^53^ These differences may render persons with these conditions more vulnerable to the negative cognitive impacts of AC medications. High AC medication exposure is also attested in adults with schizophrenia, a clinical population that, like autistic adults, has both underlying cognitive vulnerabilities such as executive functioning challenges and elevated and early exposure to AC medications. Among adults with schizophrenia, AC medications are associated with poorer verbal memory, working memory, and attention, even after controlling for current symptoms,^54,55^ and discontinuation of AC medications has been associated with improvements in cognition.^56^

Our findings suggest that, as in the general population, medications with AC effects may constitute one potential modifiable risk factor for negative aging-related outcomes among persons with autism and/or ID. Therefore, periodic reevaluation of medication regimens over time, particularly if an individual is experiencing changes in cognition, with reference to standards established for prescribing medications with high AC potency in the general population,^57^ as well as deprescribing practices^58^ may be helpful in this population.

### Limitations

We endeavored to control for indication bias by running sensitivity analyses restricting the samples to persons not prescribed antidepressant and/or antipsychotic medication(s) with AC effects. Nevertheless, possible influences of indication bias cannot be completely eliminated. Medicare Part D and Medicaid pharmacy claims captured reimbursed prescription fills but cannot establish whether medications were taken as directed and may not have captured prescriptions obtained outside of these programs. Additionally, the representation of over-the-counter medications is considerably more limited in the claims data presented here. Thus, the prevalence of AC medication exposure may have been underestimated. The resulting exposure misclassification may also have biased the estimated associations, although the direction of that bias cannot be determined.

Some classes of medications, for instance antihypertensive agents, may have protective effects in risk for vascular dementia.^59^ The examination of specific classes of medications with AC effects as these relate to the outcomes of interest is an important next step. Although we did not examine medication classes, one type of antihypertensive agent, beta-blockers (e.g., atenolol), was common across the four diagnostic groups; however, these medications were confined to the lowest AC potency category (i.e., AC=1).

Finally, while this is a national sample of autistic adults and adults with ID, it may not be representative of all adults with autism or ID. For instance, Medicaid and/or Medicare enrollees may include an overrepresentation of individuals with relatively complex medical and support needs compared to those with private health insurance.

## Conclusions

Based on nationwide claims data, these findings provide the strongest evidence to date indicating increased exposure to medications with AC effects, and greater prevalence of ADRD, falls, and mortality associated with this exposure (including after removing persons prescribed antipsychotic or antidepressant medications) among persons with autism and/or ID compared to a control group. Future work should take steps to examine onset of AC medication use and its associations with subsequent claims for aging-related outcomes in healthcare records, including incorporating a washout period that excludes or controls for AC medication exposure occurring during a period of time prior to diagnosis of outcomes such as ADRD.

## Acknowledgements

This study was supported by the National Institute of Mental Health (NIMH) under the Award Number R01MH117653 (PI: L.L.S.). This project was also supported by the Health Resources and Services Administration (HRSA) of the US Department of Health and Human Services (HHS) under cooperative agreement UT2MC39440, Autism Intervention Research Network on Physical Health (AIR-P; PI: L.L.S.). G.A.M. acknowledges support under NIMH K01MH129622. The information, content, and/or conclusions are those of the author and should not be construed as the official position or policy of, nor should any endorsements be inferred by HRSA, HHS or the U.S. Government.

This manuscript is the result of funding in whole or in part by the National Institutes of Health (NIH). It is subject to the NIH Public Access Policy. Through acceptance of this federal funding, NIH has been given a right to make this manuscript publicly available in PubMed Central upon the Official Date of Publication, as defined by NIH.

## Author Contributions

G.A.M. and G.L.W contributed to the conception and design of the study. S.T. and L.L.S. contributed to the acquisition of data. S.T. and G.A.M. contributed to analysis of data. G.A.M., S.T., L.L.S., and G.L.W. contributed to drafting of the text. G.A.M. and S.T. contributed to preparing the figures and tables.

## Potential Conflicts of Interest

Nothing to report.

## Data Availability

Due to data use agreement requirements by the Centers for Medicare and Medicaid Services, sharing of individual data from this study is not permitted.

## Supplemental Materials

### Supplemental Methods

#### Data extraction for medications

For Medicare enrollees, medication information was extracted from “Medicare Part D Event (PDE)/Drug Characteristics”. For Medicaid enrollees, medication information was extracted from the “RX” file of T-MSIS Analytic Files (TAF). Records include fee-for-service (FFS) claims, managed care encounters and supplemental payments for Medicaid, Medicaid-expansion CHIP and separate CHIP. For dually enrolled individuals, medications were identified using all claims in both Medicare and Medicaid. See Table S1. Exposure to medications with AC effects was determined based on medications recorded over the study period (2017-2019).

#### Medication routes excluded in AC potency coding

Following prior work^1,2^ and in order to be maximally conservative, in considering AC medications, those with topical, ophthalmic, or nasal routes of administration were excluded due to possible reduced systemic absorption. See Table S2 for details concerning these excluded routes of administration.

#### Coding of Medications for AC potency

Medications recorded during the study period (2017-2019) were coded for their AC potency based on the CRIDECO Anticholinergic Load Scale (CALS).^1^ Individuals who did not have a claim for a medication with AC effects during the study period were coded as “0”. For individuals who took any medication with AC effects during the study period, coding considered the highest potency AC medication taken. For example, if a given individual took three AC medications during the study period where each medication was a different AC potency (1, 2, 3), they were coded according to the highest-potency AC medication taken (i.e., AC=3).

#### Sensitivity analyses removing persons prescribed antidepressant or antipsychotic medications with AC effects

Thus, to help ameliorate concerns regarding potential indication bias, sensitivity analyses using Modified Poisson regressions excluded persons for whom antidepressant and/or antipsychotic medication(s) were prescribed. Table S3 indicates those medications considered to be exclusionary in these sensitivity analyses. The final prevalence sample for the sensitivity analyses was comprised of *N*=44,622,264 persons: autism-only, *N*=20,176; autism+ ID: *N*=20,780; ID-only: *N*=227,703; Control: *N=*44,353,605. Demographic information for each group for the sensitivity analyses sample is reported in Table S4. Results of analyses are reported in Table S5 and S6.

**Table S1.** ICD-10 Codes and/or Claims/Indicators for Variables.

| Diagnosis | Claim(s)/Indicator(s) | ICD-10 |
|---|---|---|
| Autism | 1 inpatient claim or 2 non-inpatient claims | F84.x |
| Intellectual disability | 1 inpatient claim or 2 non-inpatient claims | F70-F79 |
| Alzheimer's Disease and Related Disorders or Senile Dementia | 1 inpatient or 1 non-inpatient claim | F01.50, F01.51, F02.80, F02.81, F03.90, F03.91, F04, F05, F06.1, F06.8, G13.8, G30.0, G30.1, G30.8, G30.9, G31.01, G31.09, G31.1, G31.2, G94, R41.81, R54 |
| Falls/Fall risk (ICD-10: Slipping, tripping, stumbling and falls) | 1 inpatient or 1 non-inpatient claim | W00.XXXX-W15.XXXX, W17.XXXX<br>V00111,V00111A,V00111D,V00111S,V00121,V00121A,V00121D,V00121S,V00131,V00131A,V00131D,V00131S,V00141,V00141A,V00141D,V00141S,V00151,V00151A,V00151D,V00151S,V00181,V00181A,V00181D,V00181S,V00211,V00211A,V00211D,V00211S,V00221,V00221A,V00221D,V00221S,V00281,V00281A,V00281D,V00281S,V00311,V00311A,V00311D,V00311S,V00321,V00321A,V00321D,V00321S,V00381,V00381A,V00381D,V00381S,V00811,V00811A,V00811D,V00811S,V00821,V00821A,V00821D,V00821S,V00831,V00831A,V00831D,V00831S,V00841,V00841A,V00841D,V00841S,V00891,V00891A,V00891D,V00891S,W16012,W16012A,W16012D,W16012S,W16022,W16022A,W16022D,W16022S,W16112,W16112A,W16112D,W16112S,W16122,W16122A,W16122D,W16122S,W16132,W16132A,W16132D,W16132S,W16212,W16212A,W16212D,W16212S,W16222,W16222A,W16222D,W16222S,W16312,W16312A,W16312D,W16312S,W16322,W16322A,W16322D,W16322S,W16332,W16332A,W16332D,W16332S,W16512,W16512A,W16512D,W16512S,W16522,W16522A,W16522D,W16522S,W16532,W16532A,W16532D,W16532S,W16612,W16612A,W16612D,W16612S,W16622,W16622A,W16622D,W16622S,W16712,W16712A,W16712D,W16712S,W16722,W16722A,W16722D,W16722S,W16812,W16812A,W16812D,W16812S,W16822,W16822A,W16822D,W16822S,W16832,W16832A,W16832D,W16832S,W181,W1811,W1811XA,W1811XD,W1811XS,W1812,W1812XA,W1812XD,W1812XS,W182,W182XXA,W182XXD,W182XXS,W183,W1830,W1830XA,W1830XD,W1830XS,W1831,W1831XA,W1831XD,W1831XS,W1839,W1839XA,W1839XD,W1839XS,W19,W19XXXA,W19XXXD,W19XXXS |
| Mortality | Medicare: The combination use of Date of Death (BENE_DEATH_DT) and Valid Date of Death Switch (VALID_DEATH_DT_SW);<br>In Medicaid data: The combination use of Medicaid Death Date (EL_DOD), Medicare Death Date (MDCR_DOD), Medicare Death Day Switch (MDCR_DEATH_DAY_SW).<br>In Medicaid TAF data: The combination use of Death Date (DEATH_DT) and Indicator that Beneficiary Died During the Calendar Year (DEATH_IND);<br>For those dually enrolled and have information in both data sources, both Medicaid and Medicare data were used, and mortality in both sources will be counted. | NA |

Note. "Claims" refers to claims in Medicare and Medicaid combined. ICD-10=International Classification of Diseases, 10th Revision

**Table S2.** Medication Routes Excluded from Analyses.

| Source | Variable | Excluded |
|---|---|---|
| Codebook for the T-MSIS Analytic File (TAF) Claims | category_name | miscellaneous topical agents; ophthalmic anti-infectives; ophthalmic anti-inflammatory agents; ophthalmic antihistamines and decongestants; ophthalmic steroids with anti-infectives; topical antihistamines; topical steroids with anti-infectives |
| T-MSIS Analytic File (TAF) Claims | DOSAGE_FORM_CD | 02=Ointment;<br>03=Cream;<br>13=Lotion;<br>14=Shampoo |
| Search of NDC via RxMix version 3.2.157 ( https://mor.nlm.nih.gov/RxMix/) using the following search: rxnorm.findRxcuiById (idtype:"NDC", allsrc:"0"):idGroup.rxnormId<br>rxterms.getAllRxTermInfo | F2_route | Ophthalmic; Topical; Nasal |

**Table S3.**
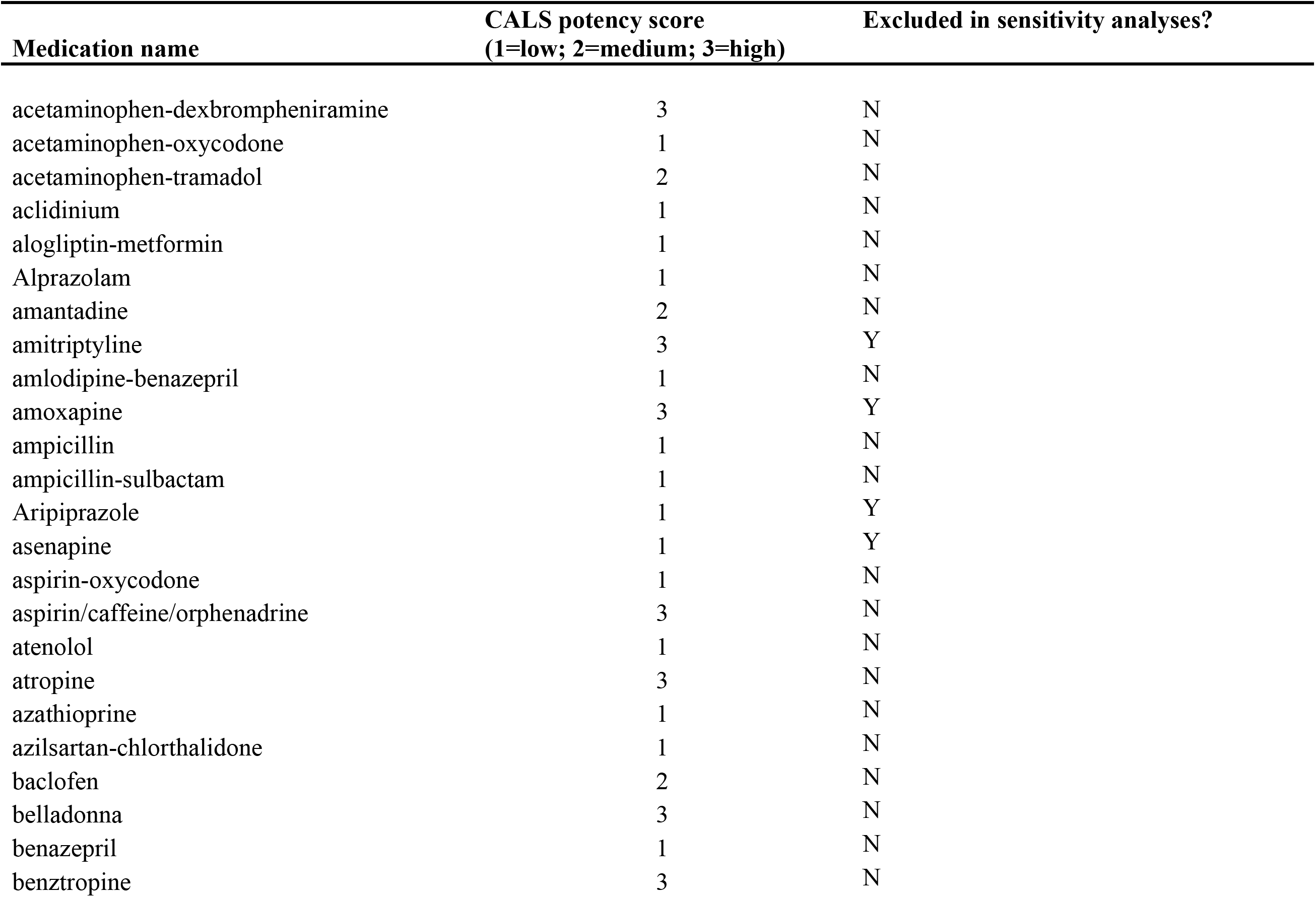
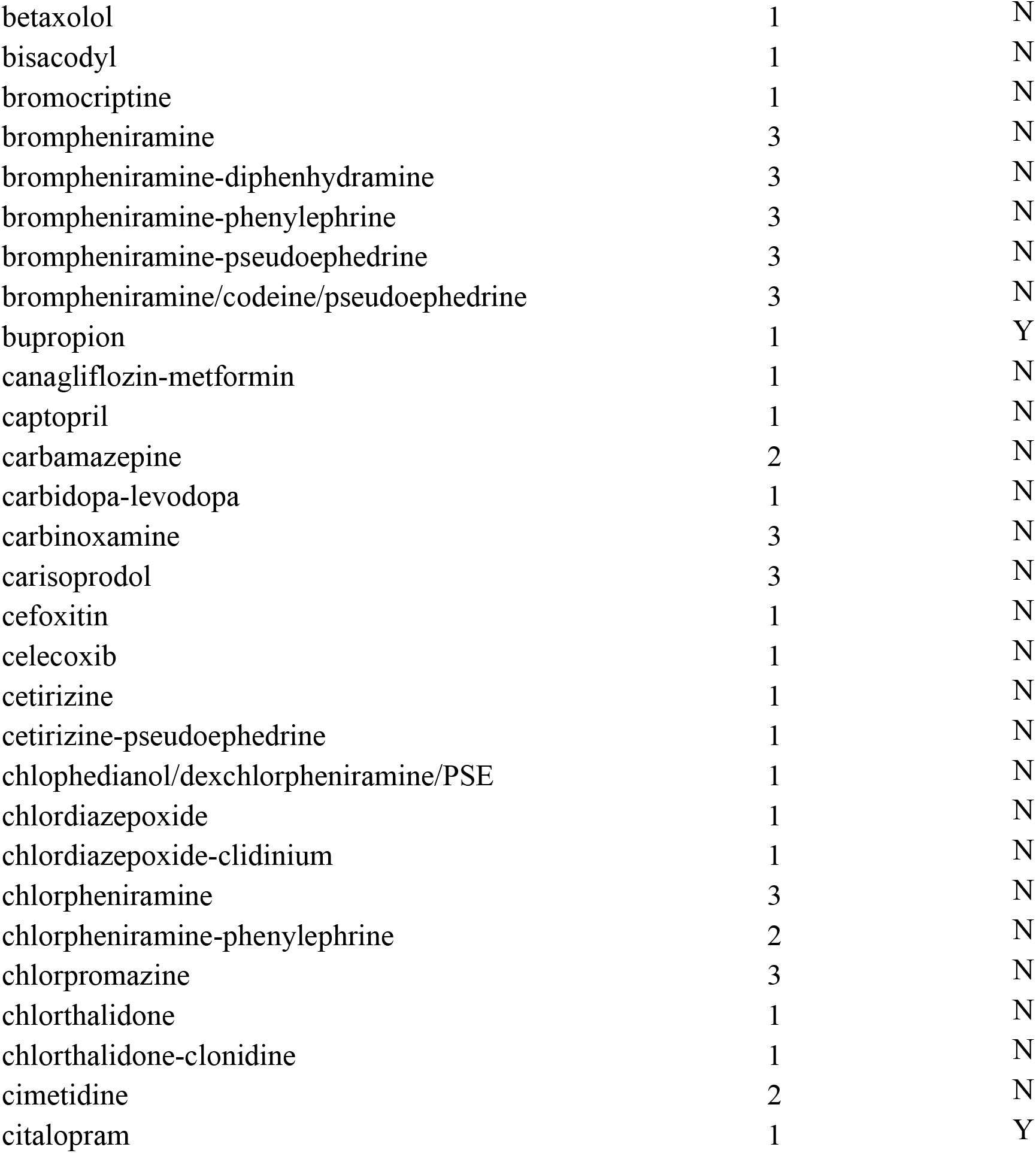
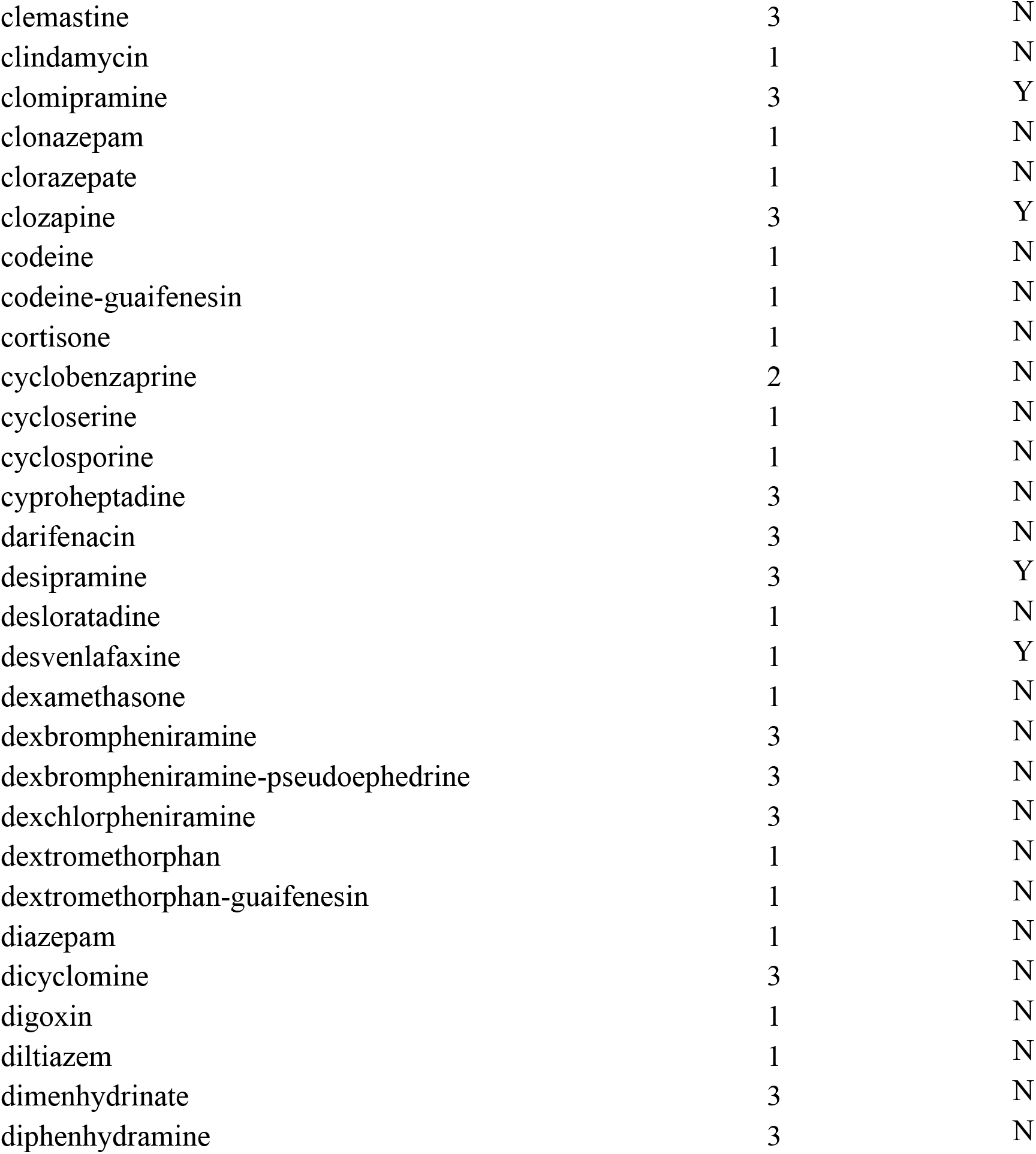
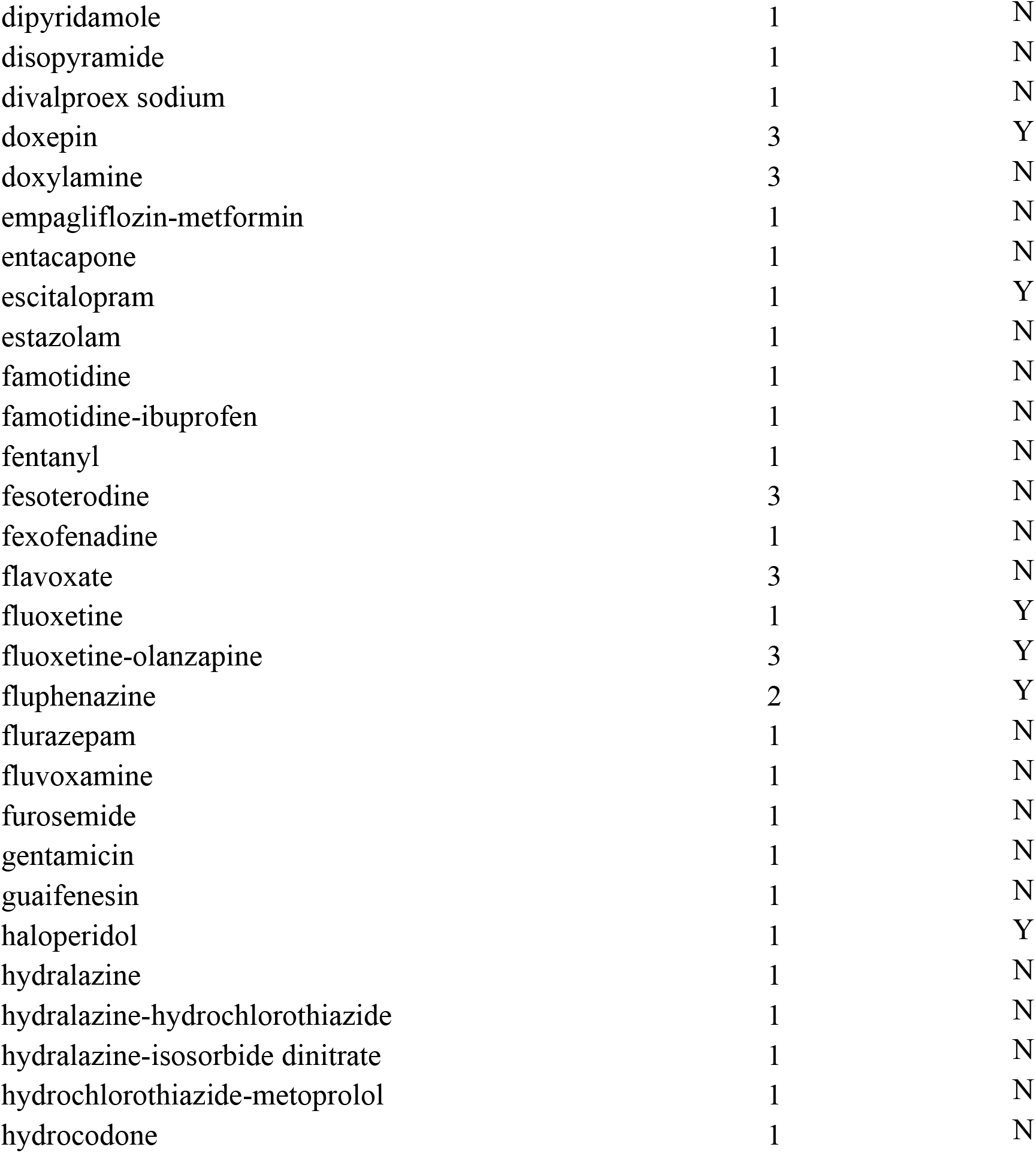
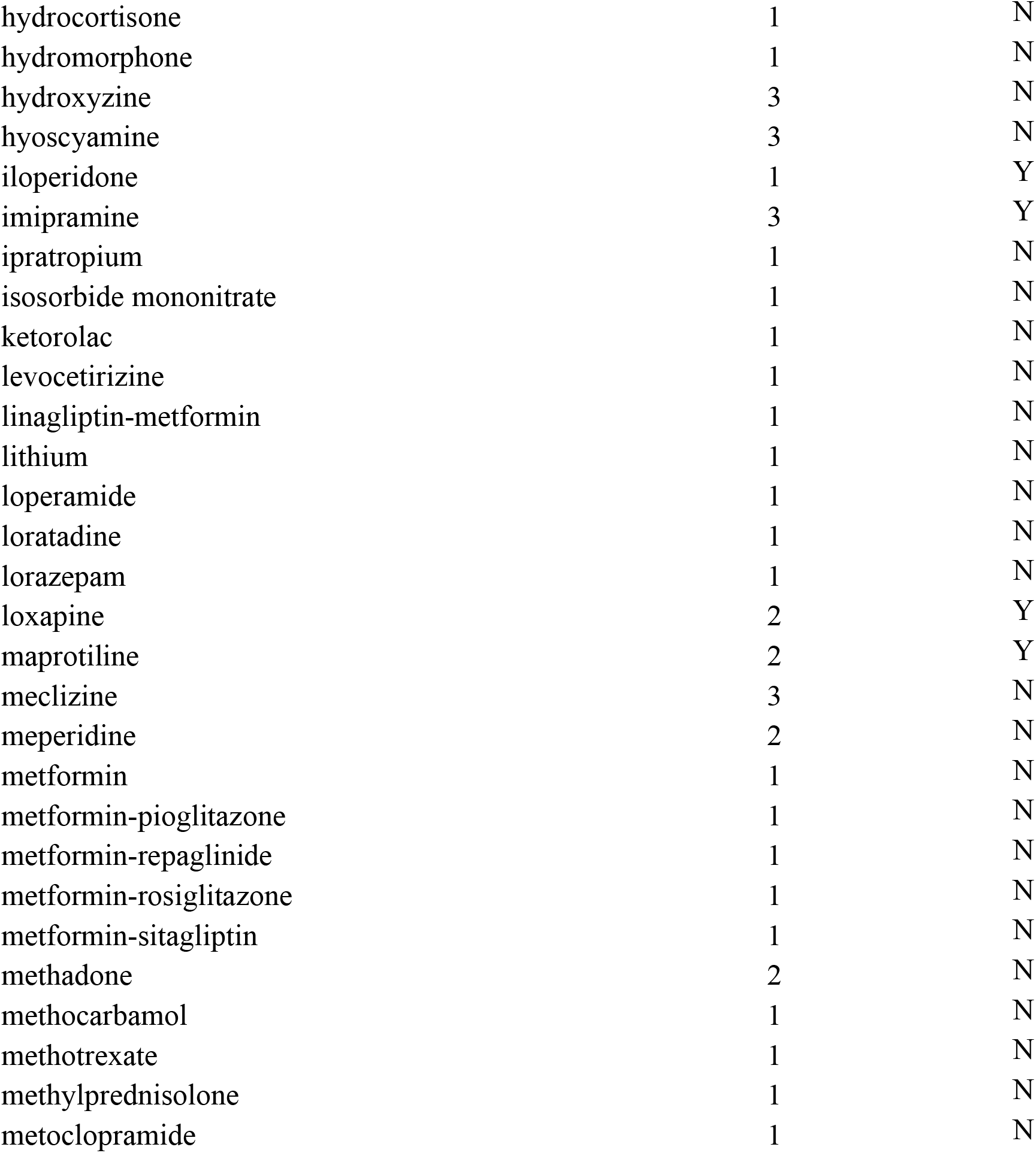
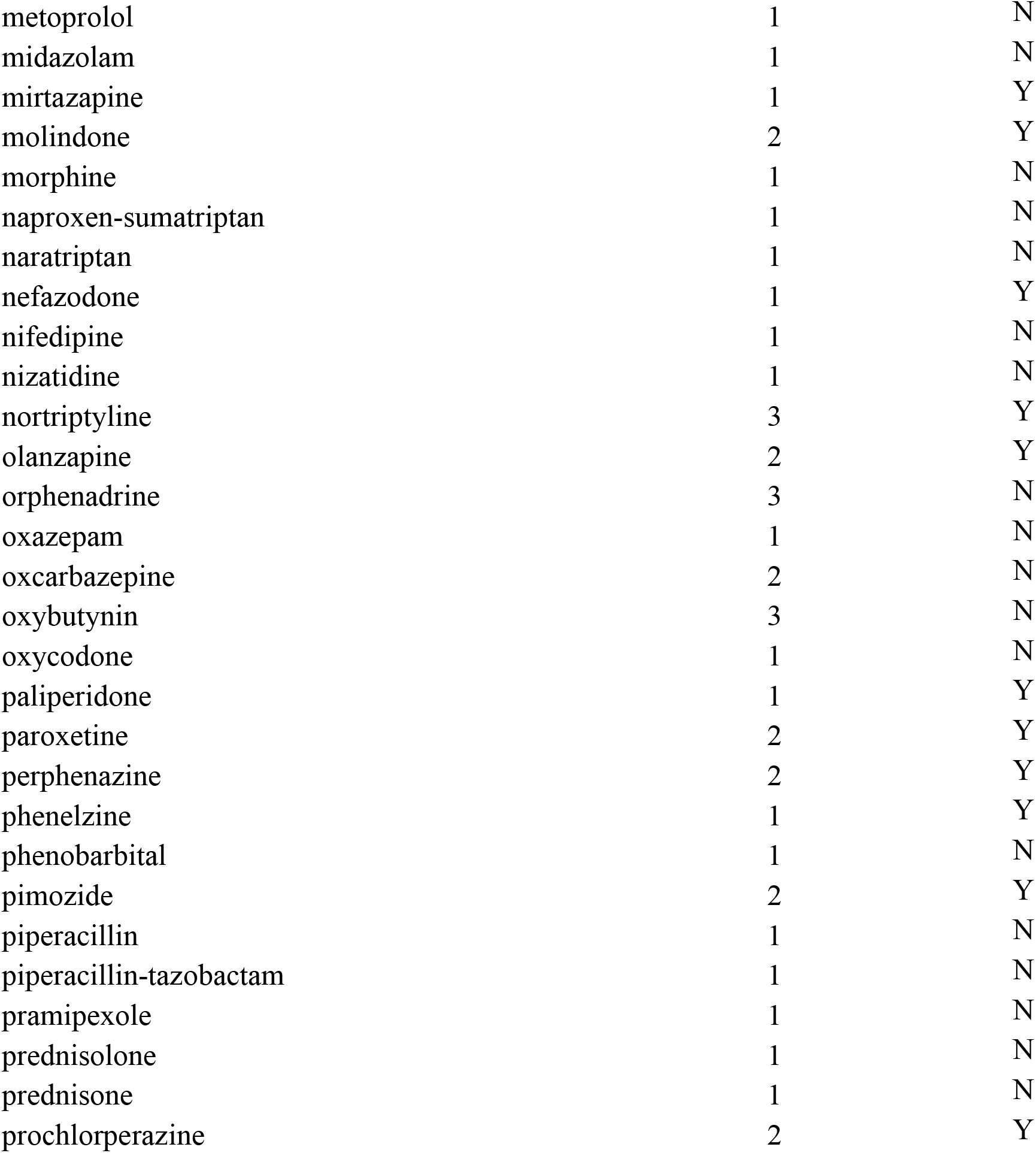
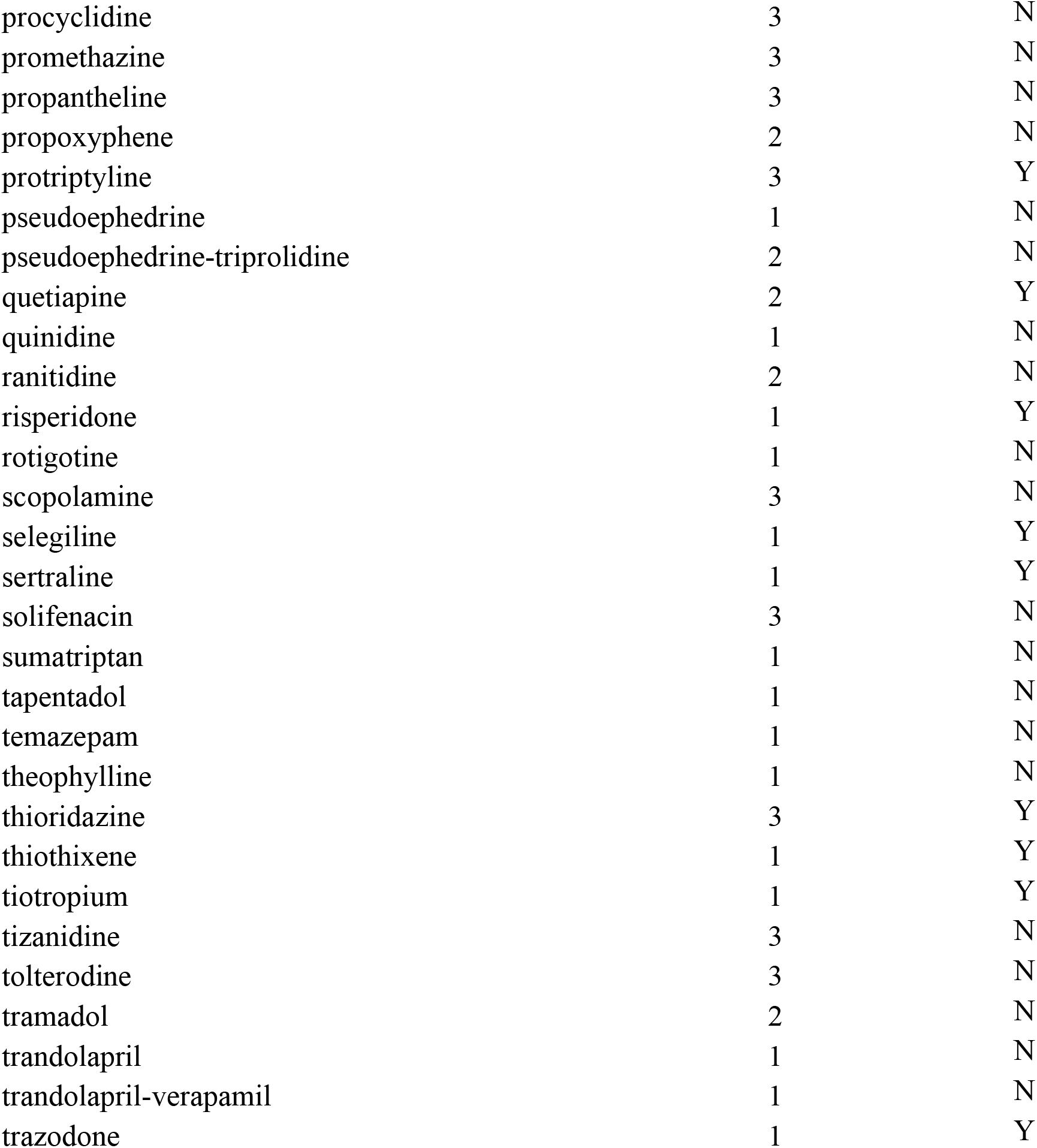
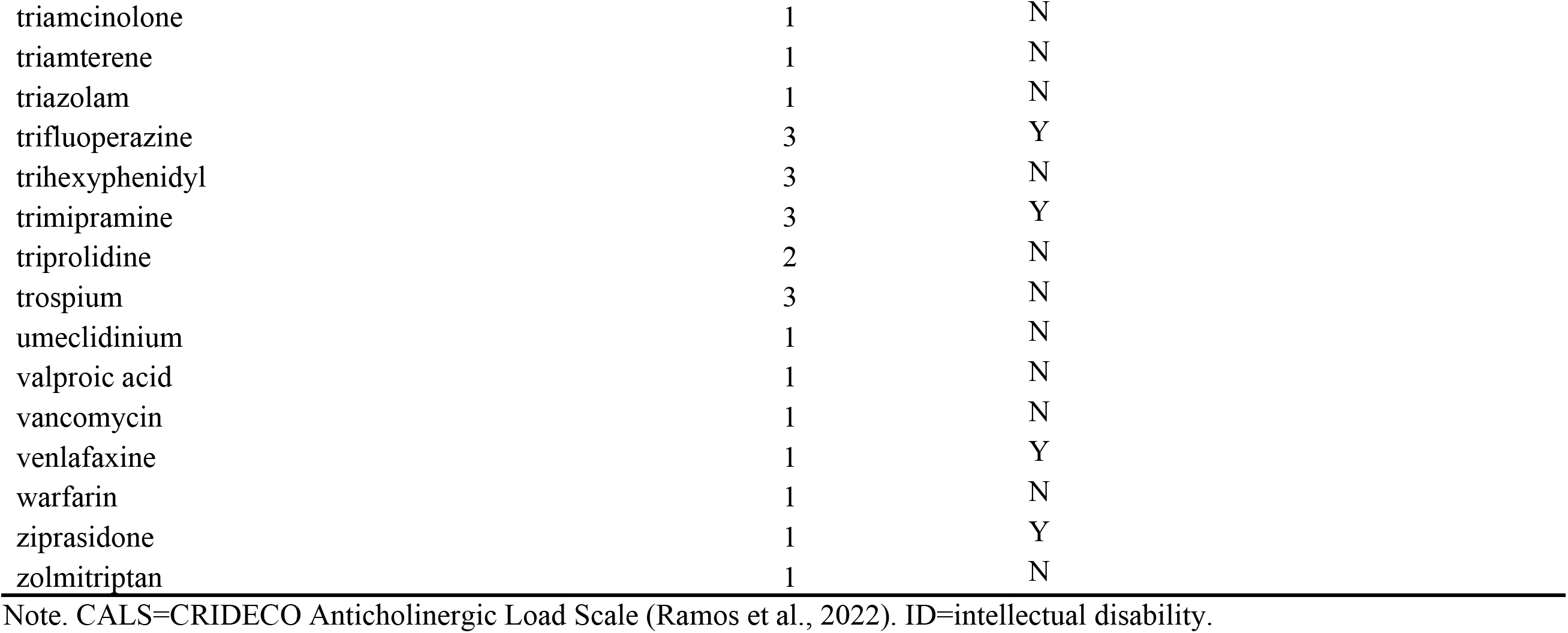
Complete List of Medications with Anticholinergic (AC) Effects Listed in Medicaid and/or Medicare Databases for Individuals in the Four Groups (Autism-only, Autism+ID, ID-only, Control) Aged 40+ Years and Potency Score According to the CRIDECO Anticholinergic Load Scale (CALS).

| Medication name | CALS potency score<br>(1=low; 2=medium; 3=high) | Excluded in sensitivity analyses? |
|---|---|---|
| acetaminophen-dexbrompheniramine | 3 | N |
| acetaminophen-oxycodone | 1 | N |
| acetaminophen-tramadol | 2 | N |
| aclidinium | 1 | N |
| alogliptin-metformin | 1 | N |
| Alprazolam | 1 | N |
| amantadine | 2 | N |
| amitriptyline | 3 | Y |
| amlodipine-benazepril | 1 | N |
| amoxapine | 3 | Y |
| ampicillin | 1 | N |
| ampicillin-sulbactam | 1 | N |
| Aripiprazole | 1 | Y |
| asenapine | 1 | Y |
| aspirin-oxycodone | 1 | N |
| aspirin/caffeine/orphenadrine | 3 | N |
| atenolol | 1 | N |
| atropine | 3 | N |
| azathioprine | 1 | N |
| azilsartan-chlorthalidone | 1 | N |
| baclofen | 2 | N |
| belladonna | 3 | N |
| benazepril | 1 | N |
| benztropine | 3 | N |
| betaxolol | 1 | N |
| bisacodyl | 1 | N |
| bromocriptine | 1 | N |
| brompheniramine | 3 | N |
| brompheniramine-diphenhydramine | 3 | N |
| brompheniramine-phenylephrine | 3 | N |
| brompheniramine-pseudoephedrine | 3 | N |
| brompheniramine/codeine/pseudoephedrine | 3 | N |
| bupropion | 1 | Y |
| canagliflozin-metformin | 1 | N |
| captopril | 1 | N |
| carbamazepine | 2 | N |
| carbidopa-levodopa | 1 | N |
| carbinoxamine | 3 | N |
| carisoprodol | 3 | N |
| cefoxitin | 1 | N |
| celecoxib | 1 | N |
| cetirizine | 1 | N |
| cetirizine-pseudoephedrine | 1 | N |
| chlophedianol/dexchlorpheniramine/PSE | 1 | N |
| chlordiazepoxide | 1 | N |
| chlordiazepoxide-clidinium | 1 | N |
| chlorpheniramine | 3 | N |
| chlorpheniramine-phenylephrine | 2 | N |
| chlorpromazine | 3 | N |
| chlorthalidone | 1 | N |
| chlorthalidone-clonidine | 1 | N |
| cimetidine | 2 | N |
| citalopram | 1 | Y |
| clemastine | 3 | N |
| clindamycin | 1 | N |
| clomipramine | 3 | Y |
| clonazepam | 1 | N |
| clorazepate | 1 | N |
| clozapine | 3 | Y |
| codeine | 1 | N |
| codeine-guaifenesin | 1 | N |
| cortisone | 1 | N |
| cyclobenzaprine | 2 | N |
| cycloserine | 1 | N |
| cyclosporine | 1 | N |
| cyproheptadine | 3 | N |
| darifenacin | 3 | N |
| desipramine | 3 | Y |
| desloratadine | 1 | N |
| desvenlafaxine | 1 | Y |
| dexamethasone | 1 | N |
| dexbrompheniramine | 3 | N |
| dexbrompheniramine-pseudoephedrine | 3 | N |
| dexchlorpheniramine | 3 | N |
| dextromethorphan | 1 | N |
| dextromethorphan-guaifenesin | 1 | N |
| diazepam | 1 | N |
| dicyclomine | 3 | N |
| digoxin | 1 | N |
| diltiazem | 1 | N |
| dimenhydrinate | 3 | N |
| diphenhydramine | 3 | N |
| dipyridamole | 1 | N |
| disopyramide | 1 | N |
| divalproex sodium | 1 | N |
| doxepin | 3 | Y |
| doxylamine | 3 | N |
| empagliflozin-metformin | 1 | N |
| entacapone | 1 | N |
| escitalopram | 1 | Y |
| estazolam | 1 | N |
| famotidine | 1 | N |
| famotidine-ibuprofen | 1 | N |
| fentanyl | 1 | N |
| fesoterodine | 3 | N |
| fexofenadine | 1 | N |
| flavoxate | 3 | N |
| fluoxetine | 1 | Y |
| fluoxetine-olanzapine | 3 | Y |
| fluphenazine | 2 | Y |
| flurazepam | 1 | N |
| fluvoxamine | 1 | N |
| furosemide | 1 | N |
| gentamicin | 1 | N |
| guaifenesin | 1 | N |
| haloperidol | 1 | Y |
| hydralazine | 1 | N |
| hydralazine-hydrochlorothiazide | 1 | N |
| hydralazine-isosorbide dinitrate | 1 | N |
| hydrochlorothiazide-metoprolol | 1 | N |
| hydrocodone | 1 | N |
| hydrocortisone | 1 | N |
| hydromorphone | 1 | N |
| hydroxyzine | 3 | N |
| hyoscyamine | 3 | N |
| iloperidone | 1 | Y |
| imipramine | 3 | Y |
| ipratropium | 1 | N |
| isosorbide mononitrate | 1 | N |
| ketorolac | 1 | N |
| levocetirizine | 1 | N |
| linagliptin-metformin | 1 | N |
| lithium | 1 | N |
| loperamide | 1 | N |
| loratadine | 1 | N |
| lorazepam | 1 | N |
| loxapine | 2 | Y |
| maprotiline | 2 | Y |
| meclizine | 3 | N |
| meperidine | 2 | N |
| metformin | 1 | N |
| metformin-pioglitazone | 1 | N |
| metformin-repaglinide | 1 | N |
| metformin-rosiglitazone | 1 | N |
| metformin-sitagliptin | 1 | N |
| methadone | 2 | N |
| methocarbamol | 1 | N |
| methotrexate | 1 | N |
| methylprednisolone | 1 | N |
| metoclopramide | 1 | N |
| metoprolol | 1 | N |
| midazolam | 1 | N |
| mirtazapine | 1 | Y |
| molindone | 2 | Y |
| morphine | 1 | N |
| naproxen-sumatriptan | 1 | N |
| naratriptan | 1 | N |
| nefazodone | 1 | Y |
| nifedipine | 1 | N |
| nizatidine | 1 | N |
| nortriptyline | 3 | Y |
| olanzapine | 2 | Y |
| orphenadrine | 3 | N |
| oxazepam | 1 | N |
| oxcarbazepine | 2 | N |
| oxybutynin | 3 | N |
| oxycodone | 1 | N |
| paliperidone | 1 | Y |
| paroxetine | 2 | Y |
| perphenazine | 2 | Y |
| phenelzine | 1 | Y |
| phenobarbital | 1 | N |
| pimozide | 2 | Y |
| piperacillin | 1 | N |
| piperacillin-tazobactam | 1 | N |
| pramipexole | 1 | N |
| prednisolone | 1 | N |
| prednisone | 1 | N |
| prochlorperazine | 2 | Y |
| procyclidine | 3 | N |
| promethazine | 3 | N |
| propantheline | 3 | N |
| propoxyphene | 2 | N |
| protriptyline | 3 | Y |
| pseudoephedrine | 1 | N |
| pseudoephedrine-triprolidine | 2 | N |
| quetiapine | 2 | Y |
| quinidine | 1 | N |
| ranitidine | 2 | N |
| risperidone | 1 | Y |
| rotigotine | 1 | N |
| scopolamine | 3 | N |
| selegiline | 1 | Y |
| sertraline | 1 | Y |
| solifenacin | 3 | N |
| sumatriptan | 1 | N |
| tapentadol | 1 | N |
| temazepam | 1 | N |
| theophylline | 1 | N |
| thioridazine | 3 | Y |
| thiothixene | 1 | Y |
| tiotropium | 1 | Y |
| tizanidine | 3 | N |
| tolterodine | 3 | N |
| tramadol | 2 | N |
| trandolapril | 1 | N |
| trandolapril-verapamil | 1 | N |
| trazodone | 1 | Y |
| triamcinolone | 1 | N |
| triamterene | 1 | N |
| triazolam | 1 | N |
| trifluoperazine | 3 | Y |
| trihexyphenidyl | 3 | N |
| trimipramine | 3 | Y |
| triprolidine | 2 | N |
| trospium | 3 | N |
| umeclidinium | 1 | N |
| valproic acid | 1 | N |
| vancomycin | 1 | N |
| venlafaxine | 1 | Y |
| warfarin | 1 | N |
| ziprasidone | 1 | Y |
| zolmitriptan | 1 | N |

Note. CALS=CRIDECO Anticholinergic Load Scale (Ramos et al., 2022). ID=intellectual disability.

**Figure S1.**
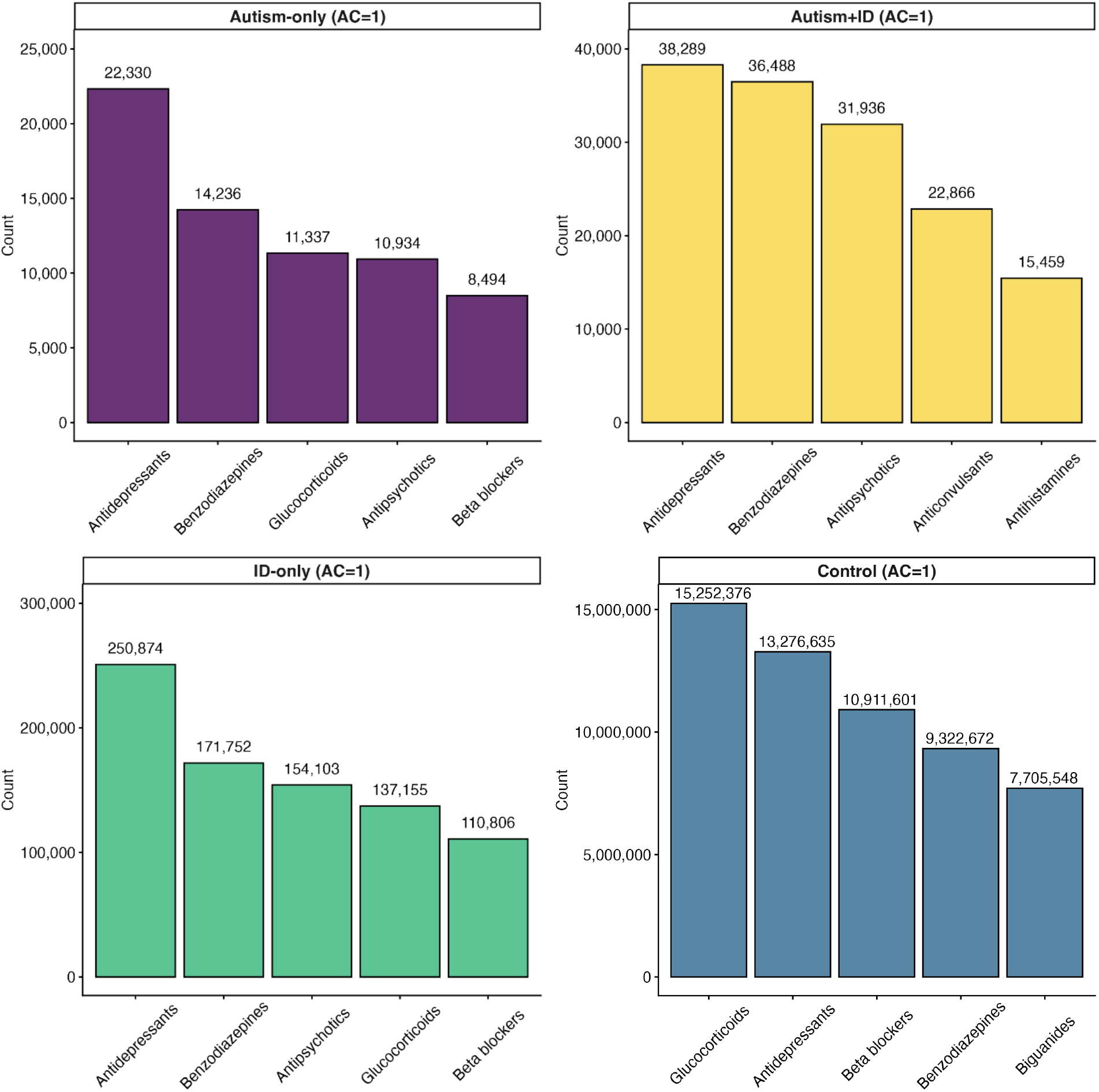
Visual depiction of the most frequent classes of anticholinergic (AC) medications with an AC potency of 1 based on the CRIDECO Anticholinergic Load Scale (CALS; Ramos et al., 2022) within each group (from top left to bottom right: autism-only, autism+intellectual disability (ID), ID-only, control).

**Figure S2.**
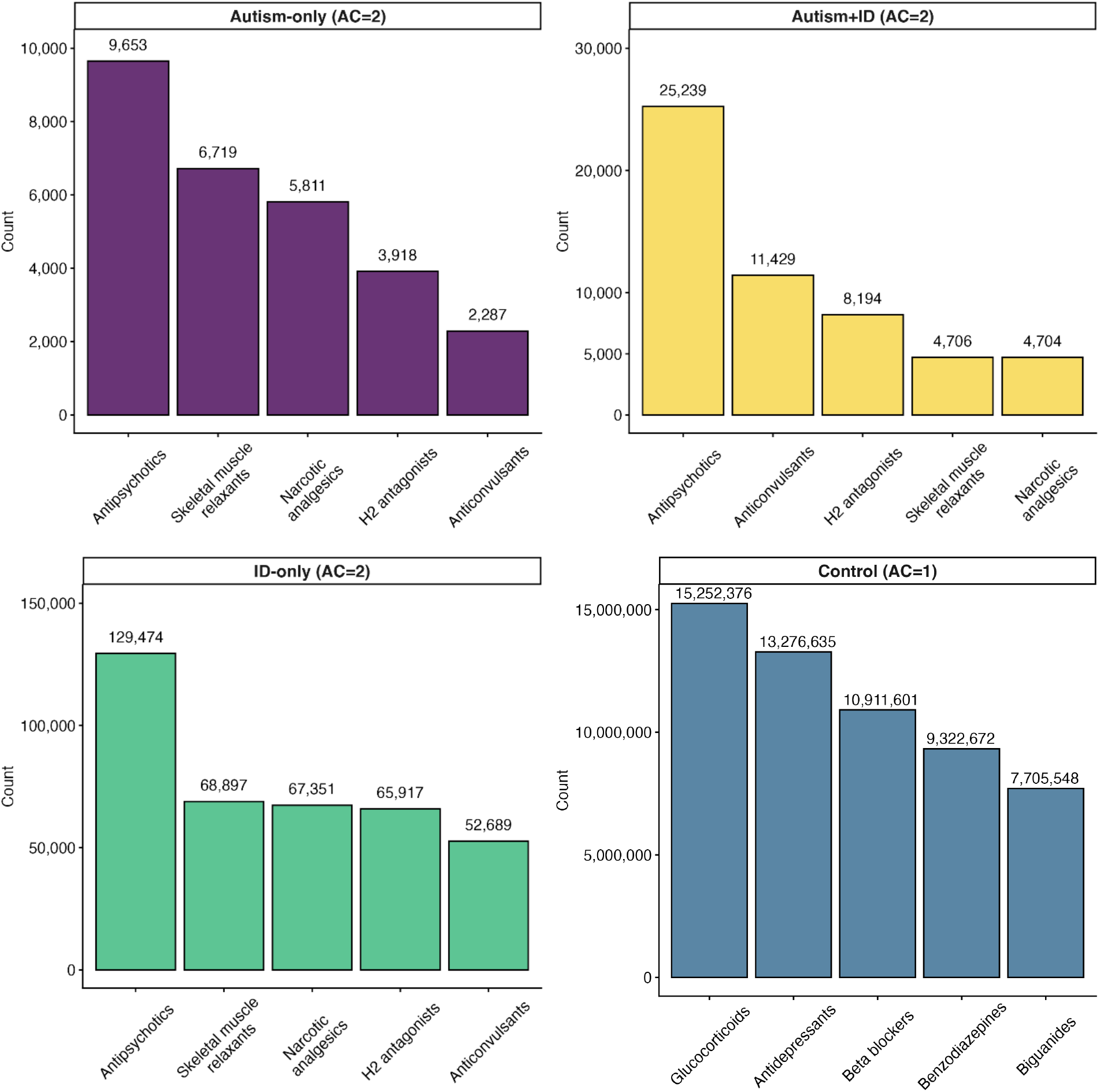
Visual depiction of the most frequent classes of anticholinergic (AC) medications with an AC potency of 2 based on the CRIDECO Anticholinergic Load Scale (CALS; Ramos et al., 2022) within each group (from top left to bottom right: autism-only, autism+intellectual disability (ID), ID-only, control).

**Figure S3.**
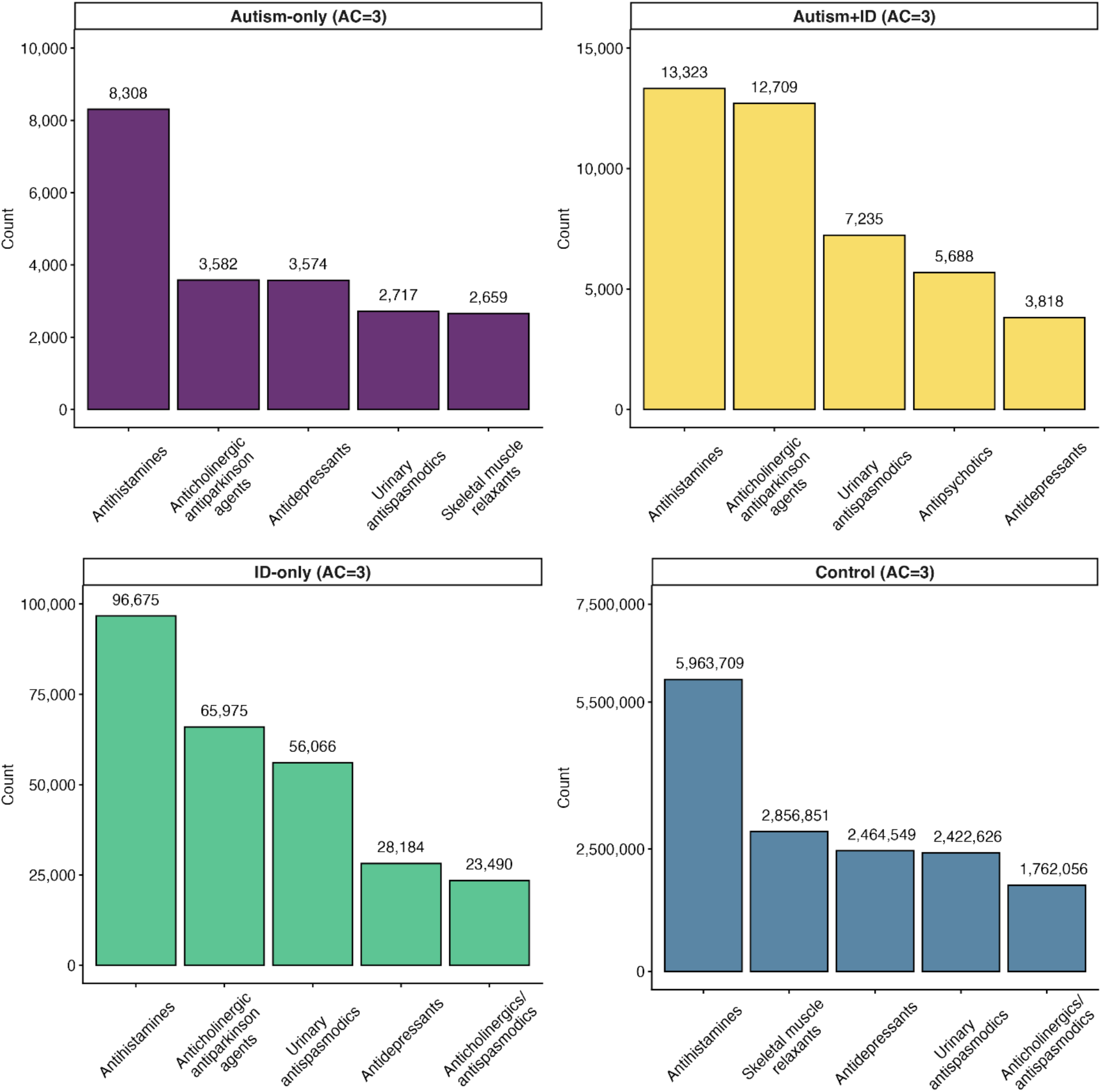
Visual depiction of the most frequent classes of anticholinergic (AC) medications with an AC potency of 3 based on the CRIDECO Anticholinergic Load Scale (CALS; Ramos et al., 2022) within each group (from top left to bottom right: autism-only, autism+intellectual disability (ID), ID-only, control).

**Table S4.** Characteristics of participants in the four groups, after removing persons prescribed antidepressant or antipsychotic medications with anticholinergic (AC) effects.

|  | **Autism only**<br>*N*=20,176 |  | **Autism + ID**<br>*N*=20,780 |  | **ID only**<br>*N*=227,703 |  | **Control**<br>*N*=44,353,605 |  |
|---|---|---|---|---|---|---|---|---|
|  | **No AC Meds** | **AC Meds** | **No AC Meds** | **AC Meds** | **No AC Meds** | **AC Meds** | **No AC Meds** | **AC Meds** |
| ***n*, (%)** | 9,755 (48.35) | 10,421 (51.65) | 6,053 (29.13) | 14,727 (70.87) | 65,495 (28.76) | 162,208 (71.24) | 23,411,587 (52.78) | 20,942,018 (47.22) |
| **Age, years** |  |  |  |  |  |  |  |  |
| Mean (SD) | 55.91 (12.95) | 56.42 (12.98) | 50.46 (9.61) | 53.10 (10.42) | 54.58 (11.85) | 56.63 (11.83) | 65.37 (12.59) | 65.83 (12.58) |
| **Age Group, *n* (%)** |  |  |  |  |  |  |  |  |
| 40-44 | 2,471 (25.33) | 2,411 (23.14) | 2,089 (34.51) | 3,801 (25.81) | 16,220 (24.77) | 29,373 (18.11) | 2,081,112 (8.89) | 1,553,719 (7.42) |
| 45-49 | 1,316 (13.49) | 1,407 (13.50) | 1,161 (19.18) | 2,443 (16.59) | 9,841 (15.03) | 22,101 (13.63) | 1,313,388 (5.61) | 1,221,786 (5.83) |
| 50-54 | 1,288 (13.20) | 1,383 (13.27) | 1,002 (16.55) | 2,481 (16.85) | 9,864 (15.06) | 24,764 (15.27) | 1,231,754 (5.26) | 1,407,867 (6.72) |
| 55-59 | 964 (9.88) | 1,246 (11.96) | 713 (11.78) | 2,129 (14.46) | 9,097 (13.89) | 24,740 (15.25) | 1,284,103 (5.48) | 1,602,840 (7.65) |
| 60-64 | 1,025 (10.51) | 1,204 (11.55) | 511 (8.44) | 1,658 (11.26) | 7,263 (11.09) | 20,845 (12.85) | 3,756,502 (16.05) | 2,788,681 (13.32) |
| 65-69 | 1,113 (11.41) | 980 (9.40) | 318 (5.25) | 1,077 (7.31) | 5,467 (8.35) | 16,261 (10.02) | 5,773,917 (24.66) | 4,211,418 (20.11) |
| 70-79 | 1,103 (11.31) | 1,187 (11.39) | 207 (3.42) | 936 (6.36) | 5,383 (8.22) | 17,381 (10.72) | 5,225,278 (22.32) | 5,345,082 (25.52) |
| 80+ | 475 (4.87) | 603 (5.79) | 52 (0.86) | 202 (1.37) | 2,360 (3.60) | 6,743 (4.16) | 2,745,533 (11.73) | 2,810,625 (13.42) |
| **Sex, *n* (%)** |  |  |  |  |  |  |  |  |
| Female | 2,572 (26.37) | 3,563 (34.19) | 1,623 (26.81) | 4,888 (33.19) | 26,257 (40.09) | 73,551 (45.34) | 11,154,046 (47.64) | 11,289,199 (53.91) |
| Male | 7,183 (73.63) | 6,858 (65.81) | 4,430 (73.19) | 9,839 (66.81) | 39,238 (59.91) | 88,657 (54.66) | 12,257,541 (52.36) | 9,652,819 (46.09) |
| **Race/Ethnicity, *n* (%)** |  |  |  |  |  |  |  |  |
| Asian/Pacific Islander | 198 (2.03) | 304 (2.92) | 139 (2.30) | 367 (2.49) | 1,855 (2.83) | 4,283 (2.64) | 1,063,668 (4.54) | 1,290,444 (6.16) |
| Black | 1,060 (10.87) | 1,318 (12.65) | 1,415 (23.38) | 3,157 (21.44) | 15,374 (23.47) | 36,242 (22.34) | 2,576,968 (11.01) | 2,976,279 (14.21) |
| Hispanic/Latino | 436 (4.47) | 715 (6.86) | 413 (6.82) | 981 (6.66) | 5,801 (8.86) | 14,806 (9.13) | 2,700,173 (11.53) | 2,566,188 (12.25) |
| Native American/Alaska Native | 60 (0.62) | 86 (0.83) | 34 (0.56) | 94 (0.64) | 747 (1.14) | 1,623 (1.00) | 179,237 (0.77) | 147,665 (0.71) |
| Multiracial/Other | 57 (0.58) | 37 (0.36) | 15 (0.25) | 18 (0.12) | 185 (0.28) | 252 (0.16) | 180,522 (0.77) | 101,512 (0.48) |
| Missing | 319 (3.27) | 256 (2.46) | 67 (1.11) | 130 (0.88) | 936 (1.43) | 1,937 (1.19) | 1,267,670 (5.41) | 789,060 (3.77) |
| White | 7,625 (78.17) | 7,705 (73.94) | 3,970 (65.59) | 9,980 (67.77) | 40,597 (61.98) | 103,065 (63.54) | 15,443,349 (65.96) | 13,070,870 (62.41) |
| **Enrolled months** |  |  |  |  |  |  |  |  |
| Mean (SD) | 32.48 (6.79) | 33.22 (6.08) | 33.35 (6.09) | 34.15 (5.14) | 33.43 (6.00) | 34.03 (5.27) | 30.96 (7.91) | 32.80 (6.41) |
| **Enrolled months group** |  |  |  |  |  |  |  |  |
| 12-24 | 1,610 (16.50) | 1,319 (12.66) | 772 (12.75) | 1,250 (8.49) | 8,137 (12.42) | 14,858 (9.16) | 5,458,049 (23.31) | 3,007,220 (14.36) |
| 25-36 | 8,145 (83.50) | 9,102 (87.34) | 5,281 (87.25) | 13,477 (91.51) | 57,358 (87.58) | 147,350 (90.84) | 17,953,538 (76.69) | 17,934,798 (85.64) |
| **Enrollment, *n* (%)** |  |  |  |  |  |  |  |  |
| Medicaid Only | 2,018 (20.69) | 2,881 (27.65) | 914 (15.10) | 1,854 (12.59) | 11,340 (17.31) | 26,845 (16.55) | 5,027,385 (21.47) | 5,090,628 (24.31) |
| Medicare Only | 4,700 (48.18) | 1,901 (18.24) | 912 (15.07) | 360 (2.44) | 15,604 (23.82) | 8,061 (4.97) | 16,875,741 (72.08) | 10,572,537 (50.48) |
| Dually Enrolled | 3,037 (31.13) | 5,639 (54.11) | 4,227 (69.83) | 12,513 (84.97) | 38,551 (58.86) | 127,302 (78.48) | 1,508,461 (6.44) | 5,278,853 (25.21) |
| **Region, *n* (%)** |  |  |  |  |  |  |  |  |
| Midwest | 2,023 (20.74) | 2,317 (22.23) | 1,212 (20.02) | 3,241 (22.01) | 14,864 (22.69) | 39,837 (24.56) | 4,421,900 (18.89) | 4,497,167 (21.47) |
| Northeast | 2,123 (21.76) | 2,408 (23.11) | 1,676 (27.69) | 4,290 (29.13) | 14,307 (21.84) | 36,956 (22.78) | 4,418,282 (18.87) | 4,532,759 (21.64) |
| South | 2,986 (30.61) | 2,787 (26.74) | 2,146 (35.45) | 4,773 (32.41) | 24,150 (36.87) | 57,695 (35.57) | 8,238,427 (35.19) | 7,064,887 (33.74) |
| West | 2,623 (26.89) | 2,909 (27.91) | 1,019 (16.83) | 2,423 (16.45) | 12,174 (18.59) | 27,720 (17.09) | 6,332,978 (27.05) | 4,847,205 (23.15) |
| **Conditions, *n* (%)** |  |  |  |  |  |  |  |  |
| ADRD | 809 (8.29) | 1,019 (9.78) | 278 (4.59) | 1,500 (10.19) | 5,289 (8.08) | 20,676 (12.75) | 1,258,382 (5.38) | 1,344,759 (6.42) |
| Fall | 813 (8.33) | 1,263 (12.12) | 462 (7.63) | 2,581 (17.53) | 6,168 (9.42) | 26,815 (16.53) | 1,587,748 (6.78) | 1,941,765 (9.27) |
| Mortality | 379 (3.89) | 545 (5.23) | 140 (2.31) | 700 (4.75) | 2,746 (4.19) | 12,195 (7.52) | 1,084,893 (4.63) | 1,148,402 (5.48) |

Abbreviations. AC=anticholinergic; ADRD=Alzheimer disease and related dementias; Dually Enrolled=dually enrolled in Medicare and Medicaid; ID=intellectual disability.

**Table S5.** Sensitivity analysis: Prevalence ratios and 95% confidence intervals of Alzheimer disease and related dementias (ADRD), falls, and mortality for persons not prescribed medications with anticholinergic (AC) effects and persons prescribed at least one medication with AC effects for 2017-2019 Medicaid and/or Medicare enrollees aged ≥40 years in the four groups after removing individuals prescribed any antidepressant or antipsychotic medication with AC effects

|  | **Autism only** |  | **Autism + ID** |  | **ID only** |  | **Control** |
|---|---|---|---|---|---|---|---|
|  | **No AC Med** | **AC Med** | **No AC Med** | **AC Med** | **No AC Med** | **AC Med** | **AC Med** |
| **ADRD** |  |  |  |  |  |  |  |
| Adjusted model | 2.637<br>(2.48,2.81) | 2.368<br>(2.24,2.50) | 2.458<br>(2.19,2.76) | 3.638<br>(3.46,3.83) | 2.544<br>(2.48,2.61) | 2.854<br>(2.81,2.89) | 0.942<br>(0.94,0.94) |
| **Falls** |  |  |  |  |  |  |  |
| Adjusted model | 1.498<br>(1.40,1.60) | 2.163<br>(2.06,2.28) | 1.579<br>(1.45,1.72) | 3.312<br>(3.20,3.43) | 1.711<br>(1.67,1.75) | 2.808<br>(2.78,2.84) | 1.285<br>(1.28,1.29) |
| **Mortality** |  |  |  |  |  |  |  |
| Adjusted model | 1.196<br>(1.10,1.30) | 1.510<br>(1.40,1.63) | 1.125<br>(0.98,1.30) | 1.802<br>(1.69,1.92) | 1.411<br>(1.37,1.46) | 1.881<br>(1.85,1.91) | 1.150<br>(1.15,1.15) |

Note. In adjusted models, the prevalence ratios are adjusted for age group, sex, race/ethnicity, enrolled months group (12-24, 25-36), enrollment type (Medicaid only, Medicare only, dual enrollment), and state. The reference level in analyses was Controls not prescribed medication(s) with AC effects.
Abbreviations. AC=anticholinergic; ADRD=Alzheimer disease and related dementias; ID=intellectual disability.

**Table S6.** Sensitivity analysis: Prevalence ratios and 95% confidence intervals of Alzheimer disease and related dementias (ADRD), falls, and mortality for anticholinergic (AC) potency levels of low, medium, and high (AC=1, AC=2, AC=3) for persons prescribed at least one medication with AC effects for 2017-2019 Medicaid and/or Medicare enrollees aged ≥40 years in the four groups after removing individuals prescribed any antidepressant or antipsychotic medication with AC effects

|  | **Autism only** |  |  | **Autism + ID** |  |  | **ID only** |  |  | **Control** |  |
|---|---|---|---|---|---|---|---|---|---|---|---|
|  | **AC=1** | **AC=2** | **AC=3** | **AC=1** | **AC=2** | **AC=3** | **AC=1** | **AC=2** | **AC=3** | **AC=2** | **AC=3** |
| **ADRD** |  |  |  |  |  |  |  |  |  |  |  |
| Adjusted model | 2.303<br>(2.14,2.48) | 2.484<br>(2.22,2.78) | 2.903<br>(2.60,3.25) | 3.062<br>(2.85,3.29) | 4.037<br>(3.66,4.45) | 4.372<br>(3.98,4.81) | 2.622<br>(2.57,2.67) | 3.188<br>(3.11,3.27) | 3.431<br>(3.34,3.52) | 1.084<br>(1.08,1.09) | 1.153<br>(1.15,1.16) |
| **Falls** |  |  |  |  |  |  |  |  |  |  |  |
| Adjusted model | 1.491<br>(1.38,1.62) | 2.445<br>(2.23,2.68) | 2.424<br>(2.21,2.66) | 2.360<br>(2.24,2.49) | 3.461<br>(3.25,3.69) | 3.562<br>(3.34,3.80) | 2.129<br>(2.09,2.17) | 2.912<br>(2.85,2.97) | 3.014<br>(2.95,3.07) | 1.546<br>(1.54,1.55) | 1.499<br>(1.49,1.50) |
| **Mortality** |  |  |  |  |  |  |  |  |  |  |  |
| Adjusted model | 1.318<br>(1.20,1.45) | 1.290<br>(1.12,1.49) | 1.153<br>(0.97,1.37) | 1.254<br>(1.14,1.37) | 1.755<br>(1.55,1.98) | 1.716<br>(1.52,1.93) | 1.455<br>(1.42,1.49) | 1.743<br>(1.69,1.80) | 1.764<br>(1.71,1.82) | 1.063<br>(1.06,1.07) | 1.036<br>(1.03,1.04) |

Note. In adjusted models, the prevalence ratios are adjusted for age group, sex, race/ethnicity, enrolled months group (12-24, 25-36), enrollment type (Medicaid only, Medicare only, dual enrollment), and state. The reference level in analyses was Controls prescribed a low potency (AC=1) AC medication.
Abbreviations. AC=anticholinergic; ADRD=Alzheimer disease and related dementias; ID=intellectual disability.

## Notes

### Competing Interest Statement

The authors have declared no competing interest.

### Author Declarations

The use of these data for research was approved via HIPAA waiver by the institutional review board of Drexel University (protocol #2501010962).

